# Does immersive VR improve diagnostic accuracy of post-stroke spatial neglect relative to conventional and digital tests?

**DOI:** 10.1101/2025.02.24.25322775

**Authors:** Elise Palmans, Nora Tuts, Karla Michiels, Eline Note, Eva Van den Bussche, Céline R. Gillebert, Hanne Huygelier

## Abstract

**Objective:** Assessing post-stroke spatial neglect in immersive Virtual Reality (iVR) may improve diagnostic accuracy due to its ability to combine experimental control with increased ecological validity. This study investigated the diagnostic accuracy of an iVR assessment relative to conventional and non-iVR digital tests. Additionally, feasibility was evaluated.

**Method:** Stroke patients performed an iVR assessment, conventional pen-and-paper tests (3 Cancellation tests and a line bisection test), and non-iVR digital tests (Digital Cancellation and Posner test). A new Bayesian statistical method was developed to address the absence of a gold standard for neglect. Instead of conventional diagnostic cut-offs, we quantified the uncertainty of classifying patients as having neglect or not and incorporated this into estimating diagnostic accuracy. Feasibility was evaluated using recruitment and drop-out rates, sample characteristics, cybersickness and user experience.

**Results:** 54 stroke patients and 56 healthy controls completed the study. Both iVR and non-iVR digital tests had higher diagnostic accuracies than pen-and-paper tests, although differences were not statistically significant due to high diagnostic uncertainty (resulting from inconsistencies across tests). For 6 patients, the iVR assessment and Posner test indicated neglect, while pen-and-paper tests did not. The iVR assessment was feasible for most patients, with low cybersickness and positive evaluations.

**Conclusion:** The iVR assessment and Posner test identified spatial biases missed by conventional tests. Inconsistencies across tests highlight the complexity of neglect assessment, suggesting that future studies should explore crucial test characteristics needed for a precise and sensitive evaluation. Furthermore, results established good feasibility of iVR neglect assessment.

## 1. Introduction

After having a stroke, around 30% of patients are affected by an inability to orient, perceive and interact with stimuli (e.g., objects, sounds, persons) on the contralesional side of space despite having apparently normal perception (Esposito et al., 2021; Heilman et al., 1987, 2000). This neuropsychological condition is referred to as **spatial neglect** (for simplicity here referred to as “neglect”). Neglect patients often stay longer in the hospital and are less independent in their daily lives after their stroke (Chen et al., 2015; Nijboer et al., 2013).

Research indicates that neglect patients exhibit both spatial and non-spatial attention deficits (Corbetta & Shulman, 2011). Spatial deficits involve a lateralized bias in sensory processing, with sensitivity improving from contralesional to ipsilesional locations, and are linked to motor imbalances causing ipsilesional deviations in eye, head, and body orientation (Corbetta & Shulman, 2011). Studies report that left-sided neglect patients often show eye and head orientations deviating toward the ipsilesional side, especially in the acute post-stroke phase (Becker & Karnath, 2010; Fruhmann Berger et al., 2006; Schindler & Kerkhoff, 1997). Non-spatial deficits (e.g., impaired sustained attention, the ability to maintain focus over time) also play a significant role in neglect and may explain its persistence and severity (Husain & Rorden, 2003; Robertson, 2001).

Developing effective rehabilitation protocols for neglect begins with obtaining a precise and reliable assessment. While a wide range of assessment tools for neglect exist, there is still no universally accepted ‘gold standard’ (i.e., a single, validated tool that is widely recognized as the best) (Azouvi et al., 2006; Checketts et al., 2021; Evald et al., 2021; Moore et al., 2022; Pedroli et al., 2015). Clinicians commonly assess neglect informally based on spontaneous observations (i.e. without making use of standardized instruments and norms; Checketts et al., 2021). Observational scales, such as the Catherine Bergego Scale (CBS), can be used to assess typical neglect-related symptoms (e.g., eating food from one side of the plate only or bumping into objects on the neglected side of space) in a more structured way (Azouvi et al., 2003; Ten Brink et al., 2013). However, it is not always feasible for a single therapist to observe all daily activities evaluated by the CBS within one observation period or in the same observational context (Chen et al., 2012).

Next to these observations, neuropsychologists commonly use cancellation and line bisection tests to assess neglect. These pen-and-paper tests are easy-to-administer, but they are insensitive to mild forms of neglect (Azouvi et al., 2002; Bonato & Deouell, 2013; Schendel & Robertson, 2002). Indeed, concerns about the sensitivity of current neglect tests are well documented (Azouvi et al., 2006; Moore et al., 2022) and in addition, concerns about the specificity have also been raised (Huygelier, Moore, et al., 2020). In more chronic stages, neglect patients have often learned to compensate on “static” pen-and-paper tests when repeatedly administered, resulting in subtle neglect-related problems remaining undetected (e.g., not noticing objects fast enough to appropriately interact with them) (Buxbaum et al., 2004; Geiser et al., 2022; Spreij et al., 2020). Researchers have also argued that pen-and-paper tests lack generalization to the daily-life situations patients encounter when they are released from the hospital (e.g., in traffic or in a busy crowd), limiting their ecological validity (Rizzo et al., 2004; Tsirlin et al., 2009). Consequently, it is commonly believed that more subtle neglect-related problems and problems occurring in everyday life are not fully captured by pen-and-paper tests (Ogourtsova et al., 2018).

In the quest for increased sensitivity, some argue that digital tests (e.g., a Posner cueing test) provide a viable solution (Bonato & Deouell, 2013; Rengachary et al., 2009a). Digital tests can offer a more sensitive alternative for neglect assessment as they can record more information simultaneously (e.g., response times, accuracy, eye-tracking, search-strategies), and stimuli can be presented repeatedly in a standardized way (Bonato & Deouell, 2013; Kaufmann et al., 2020a, 2021). Additionally, digital tests can be made more resource-demanding by implementing several difficulty levels or adding dual task elements. Due to these properties, Bonato and Deouell (2013) argue that digital tests are more resistant to the development of compensatory strategies than pen-and-paper tests, potentially making such instruments more suitable to accurately track neglect over time. Although digital tests might offer higher sensitivity than pen-and-paper tests, their ecological validity is still limited. Digital tests on non-immersive screens typically capture a small part of the visual field, while in daily life we need to attend signals coming from the periphery as well as the central visual field. Moreover, digital tests originating from experimental psychology (such as the Posner cueing test) are often administered with a chin rest and patients are asked not to make eye movements. While such constraints maximize internal validity, they impose a scenario distinct from natural exploratory behavior in daily life where eye movements, head movements and covert attention shifts are typically coordinated (Karnath, 1998; Karnath & Niemeier, 2002).

In this context, the latest generation of commercial-grade high-fidelity head-mounted immersive Virtual Reality (iVR), such as the Meta Quest or HTC devices, are particularly compelling. Implementing iVR in the assessment of neglect has the potential to improve ecological validity, while the same advantages that digital tests offer remain present. These iVR systems provide an extensive field of view and can hereby immerse the user in a rich, multimodal, 3D world (Huygelier et al., 2020; Tsirlin et al., 2009). They allow the presentation of stimuli at different visual angles while considering the patient’s head and/or eye movements in real-time, and thus the assessment of the (visuo)spatial bias in a way that integrates experimental control (i.e., internal validity) with increased ecological validity (measuring spatial attention and exploration in a 360° and 3D environment).

Several studies have explored the added value of iVR in the assessment of neglect, showing that iVR does not necessarily result in increased sensitivity for neglect diagnosis (see Table 1). For example, both Knobel et al. (2020) and Kim et al. (2021) developed iVR Cancellation tests. Whereas Kim et al. (2021) found good agreement between neglect on the iVR Cancellation test and neglect on pen-and-paper tests, Knobel et al. (2020) found lower sensitivity for their iVR Cancellation test compared to a pen-and-paper version. Moreover, Hougaard et al. (2021) concluded that their iVR test did not pick up on neglect for all stroke patients with neglect on the Kessler Foundation Neglect Assessment (KF-NAP). Other studies did find increased diagnostic accuracy for iVR compared to conventional neglect tests (Belger et al., 2023; De Boi et al., 2024; Painter et al., 2024; Thomasson et al., 2023). As previous studies differ from each other on multiple aspects, such as the type of task (e.g., street crossing, exploring a virtual museum or a Cancellation test in VR), patient samples and reference tests, it is hard to provide an explanation for these mixed findings. In order to advance this research field, several limitations should first be addressed.

**Table 1.**
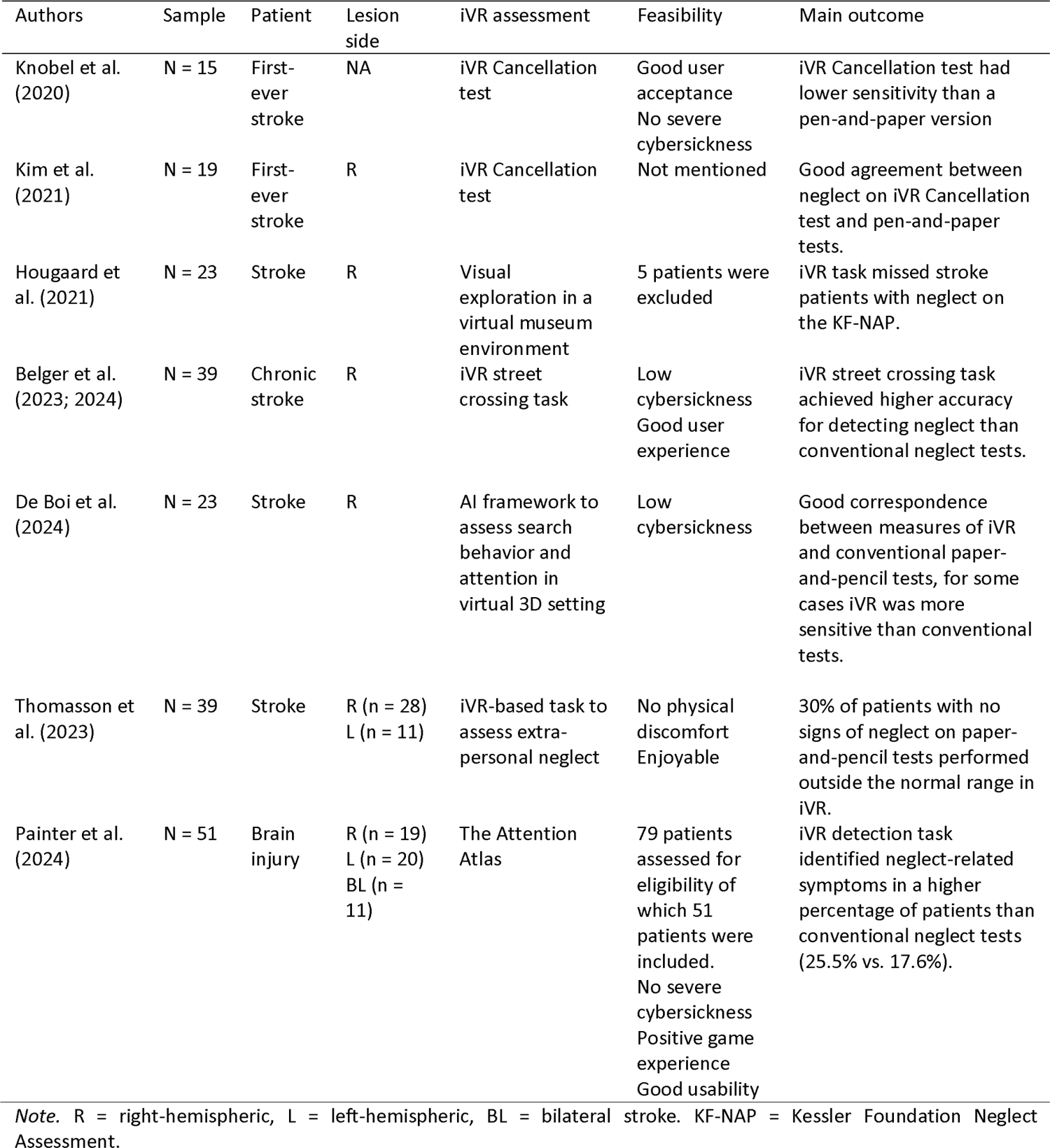
Overview of studies on iVR assessment of neglect up to 2024.

A first limitation concerns the inexistence of comparing novel iVR tools to other digital tests such as the Posner test or a Digital Cancellation test. None of the above-mentioned studies investigated whether iVR-based outcome measures could be superior relative to non-iVR *digital* tests. It thus remains open whether iVR is truly of added value for diagnostic accuracy in clinical practice. To address this, novel iVR assessments should not only be compared to conventional pen-and-paper tests, but also to other (non-iVR) digital tests that may be equally sensitive to subtle spatial biases (Rengachary et al., 2009b).

A second limitation concerns the way diagnostic accuracy of novel neglect tests is studied. That is, the standard approach to study diagnostic accuracy is to compare a novel test to a gold standard determination of “disease” status (Kohn et al., 2013). Hence, to evaluate diagnostic accuracy of novel iVR tests, the above-mentioned studies compared the iVR tool to a reference test (e.g., pen-and-paper tests). Next, indices such as sensitivity and specificity are estimated. However, this approach is sensitive to the ‘imperfect gold standard bias’, which happens when the reference test misclassifies patients. As conventional neglect tests have been reported to lack sensitivity and specificity (Azouvi et al., 2002; Huygelier, Moore, et al., 2020; Moore et al., 2022), this is a pervasive challenge in neglect research. The imperfect gold standard bias renders the estimates of the novel test’s sensitivity, specificity, or both ambiguous (Kohn et al., 2013). Kohn et al. (2013) clarify that a first step to solve this problem is to simply recognize this imperfect gold standard bias, but that there’s currently no uniform solution to this limitation. In the current study we acknowledge this by quantifying the diagnostic uncertainty of a patient’s neglect status.

A third limitation concerns the current approach to investigate the feasibility of novel iVR tests in the stroke population. Most of the above-mentioned studies, except for Painter et al. (2024), did not systematically report recruitment rates which could have led to biased results (Altman et al., 2012; Hopewell et al., 2022; Toerien et al., 2009). A novel assessment tool has clinical value only if it is feasible to implement in a representative group of its target population. Therefore, it is essential for studies to report recruitment and drop-out rates (i.e., number of exclusions, drop-outs, along with the reasons for each) and to describe the sample of patients they intended to assess with the novel tool. This transparency clarifies for which stroke patients the assessment tool is feasible and for whom it is not. Additionally, since cybersickness depends on the design of an iVR task (Jasper et al., 2023), monitoring cybersickness symptoms - such as nausea, headaches, disorientation and fatigue – is crucial for all novel iVR tools. Finally, evaluating the experience of end-users is also important, as future clinical implementation will depend on how a tool is perceived by the stakeholders.

In the current study, we investigated whether an iVR assessment, capturing spatial biases in task performance (measured in head-contingent space) and spontaneous biases in head orientation during exploration, could improve diagnostic test accuracy for post-stroke spatial neglect. Crucially, we did not only compare the iVR assessment to conventional pen-and-paper neglect tests, but also investigated whether the iVR assessment was superior relative to other non-iVR digital tests - a Posner cueing test and a Digital Cancellation test - that may be equally sensitive to subtle spatial biases (e.g., Dalmaijer et al., 2015; Gillebert et al., 2011; Rengachary et al., 2009). To study diagnostic accuracy, we developed a novel statistical approach that quantifies the uncertainty of the neglect diagnosis. Hereby, we aimed to overcome the ambiguity that is present in current diagnostic accuracy studies that compare the novel test to conventional tests with imperfect gold standard biases. Additionally, we evaluated the feasibility of the iVR assessment by assessing the following parameters: recruitment and drop-out rates, patient characteristics of the intention-to-assess sample, cybersickness symptoms pre- and post-iVR and user experience. Based on our earlier feasibility study, we expected positive user experience and less cybersickness after versus before iVR Table 1. Overview of studies on iVR assessment of neglect up to 2024. (Huygelier, Schraepen, et al., 2020).

## 2. Methods

### 2.1. Participants

Adult stroke patients residing at the rehabilitation facility of the University Hospital Leuven (Campus Pellenberg) were consecutively recruited. Patients who were able to provide informed consent were eligible to take part in the study. Severe visual (e.g., uncorrected or uncorrectable eye abnormalities), auditory (e.g. insufficiently able to perceive auditory stimuli), motor or language impairments, assessed clinically, were grounds for exclusion if they prevented the administration of the sessions. In addition, there were several exclusion criteria to use the iVR device (i.e., a cochlear implant, pacemaker, trepanation, epilepsy). Patients who had a severe psychiatric disorder, neurocognitive disorder or neurological history were also excluded. All study procedures were approved by the ethical committee of the UZ Leuven/KU Leuven (S61410 and G-2022-4775-R2(MIN)) and were in accordance with the Helsinki declaration. A total of 56 healthy controls were recruited to establish a normative range for the digital tests used in this study (see Appendix A).

### 2.2. Materials

#### 2.2.1. Immersive Virtual Reality (iVR) assessment of spatial neglect

The diagnostic module of the iVR application HEMIRehApp (Huygelier et al., 2024a) was administered to patients, using one of three head-mounted devices with minimal differences in hardware specifications (i.e., the Oculus Rift CV1, Oculus Rift S or Meta Quest 2) (Meta Quest 2, Oculus Rift S | Meta, n.d.). The three head-mounted devices have similar resolution per eye (width: 1080-1440, height: 1200-1600), refresh rates (72 - 90Hz), field of view (horizontal: 87° - 93°, vertical: 88° - 93°), weight (470g – 590g) and all devices use 6 degrees-of-freedom for tracking the position and orientation of the headset. The layout of the buttons on the handheld controllers (Oculus Touch first and second generation) are identical.

Patients were first taught how to use the iVR application in an iVR tutorial session. A series of practice trials were performed in which patients learnt to press the buttons on the handheld controller and select information in the iVR environment. Then, in the iVR assessment, participants were asked to perform a visual discrimination task in three scenes (i.e., vegetable garden, lake, forest) with three pairs of targets corresponding to each scene (i.e., ladybug-butterfly, apple-pear, duck-ducklings). There was a unique target-button combination (e.g., press “A” to catch the ladybug and press “B” to catch the butterfly). Target locations were uniformly distributed within an area subtending 30° in the left to the right side of the visual field and 5° in the upper and lower visual field. Targets were presented relative to the head midline of the patient at target onset. Targets were presented for 3 seconds or until the patient responded. The assessment was finished when a total of 225 trials (i.e., 75 trials per scene) was completed. Before each scene, patients performed 10 practice trials. On each trial, feedback was presented to indicate whether the patient’s response was correct (i.e., green checkmark), incorrect (i.e., red cross) or whether the patient did not make a response (i.e., blue exclamation mark and sound). Spontaneous biases in exploration using head movements (i.e., left- or rightward orientation of the head) were measured by sampling head orientation at 2Hz before targets were presented at the start of each scene (45s in total, 87 samples). In addition, accuracy of target discrimination was registered. Details about the iVR application are reported in (Huygelier, Schraepen, Lafosse, et al., 2020; Huygelier et al., 2024b).

#### 2.2.2. Feasibility of iVR assessment

To assess the feasibility of iVR assessment we registered the number of successfully tested patients, reasons for exclusion from iVR assessment and drop-out reasons. In addition, we assess patients’ *user experience with the iVR app,* by asking them to evaluate different aspects of the iVR tutorial and assessment using a 23-item User Experience scale (with three subscales: Experience, Presence and Usability) developed by our research team (Huygelier et al., 2019). The scale demonstrated excellent internal consistency in previous research (Huygelier et al., 2019). Each item was rated on a 5-point Likert scale ranging from totally disagree (1) to totally agree (5) with “3” as a neutral midpoint. To assess cybersickness, the Dutch Simulator Sickness Questionnaire (SSQ) was administered (Kennedy et al., 1993), which is a 16-item questionnaire that assesses common cybersickness symptoms (e.g., general discomfort, fatigue, headache, nausea). Each SSQ item was rated on a scale from ‘no discomfort’ (0) to ‘severe discomfort’ (4).

#### 2.2.3. Conventional pen-and-paper tests of spatial neglect

Four conventional pen-and-paper tests were administered on an A4 paper in landscape orientation (Checketts et al., 2020). Three Cancellation tasks were administered (Table 1). In each Cancellation task patients were instructed to cross out targets and ignore distractors spread across the page using a pencil. For each Cancellation test the difference in the number of cancelled targets for each side of the page (left and right) was used to quantify a spatial bias. In the *McIntosh line bisection task* (McIntosh, 2017; McIntosh et al., 2005), patients were presented with 32 horizontal lines depicted on an A4 page in landscape orientation. Patients were instructed to mark the middle of each line and tap the table in between each response. The task consisted of 4 types of lines that differed in line length (i.e., 4 and 8 cm) and how the left and right endpoint of the line are positioned on the page. To quantify a measure of spatial bias, the distance between the mark made by the patient and the middle of the page was measured with a ruler. Then, the influence of the position of the left and right endpoints on this distance is contrasted (i.e., endpoint weighting bias, EWB) (McIntosh et al., 2005, 2017).

The *Catherine Bergego scale* (CBS) (Azouvi et al., 2003; Ten Brink et al., 2013) is a systematic observation scale and was administered by the researchers. The CBS consists of 10 items that are rated on a scale ranging from 0 (no neglect), 1 (mild), 2 (moderate) to 3 (severe neglect) for several daily life activities (e.g., eating food, brushing hair, navigation, locating personal belongings). CBS scores are interpreted as no (0-5), mild (6-10), moderate (11-20) to severe (21-30) neglect (Ten Brink et al., 2013). Ten Brink et al. (2013) reported a moderate inter-rater agreement (ICC=.65) and moderate association with performance on neglect tests for the Dutch CBS in stroke patients.

#### 2.2.4. non-iVR digital tests of spatial neglect

We administered two non-iVR digital tests to measure spatial neglect, previously described in Huygelier et al. (2024).

First, a *Posner test* was used, this was previously suggested as one of the most sensitive neglect tests (i.e., area under the roc curve > .90 to differentiate stroke patients from healthy controls) (Rengachary et al., 2009c). In the Posner test, patients had to detect a left- or right-sided target as quickly as possible. The target (i.e., a highly salient picture) was presented after a left- or right-sided cue (i.e., white square) and was equally likely to occur at the same location as the cue (valid) or at a different location (invalid). To ensure that patients respond to target events, rather than make anticipatory responses, 20% catch trials were presented (presentation of cue, but no target). Patients completed a maximum of 400 trials (4 blocks of 100 trials each), but if they experienced fatigue the task could be prematurely ended (see Appendix A for details about task completion). All patients responded infrequently (< 12%) on catch trials (Appendix A).

Second, a *Digital Cancellation test* was administered. Patients were instructed to click with the computer mouse on 50 targets (full outlined line drawings), among 100 distractors (line drawings with an upper or lower gap) spread across the screen. Once a target had been clicked, a blue line appeared on the target to indicate that it had been crossed out. One Cancellation trial (i.e., a sheet with 150 stimuli) was presented for 4 minutes or until the participant indicated that they had finished the trial. Stimuli were placed within a horizontal angle of 11° to the left and right side and within a vertical angle of 7.3° to the upper and lower side. To ensure high precision of spatial bias measures in the Cancellation test, we administered multiple Cancellation trials (i.e., 12 trials). If patients experienced fatigue, the task could be prematurely stopped (see Appendix A). Patients could practice the Cancellation task during one practice trial. This enabled patients to familiarize themselves with the computer mouse.

#### 2.2.5. General cognitive and mood screening

A semi-structured interview was conducted to obtain demographic information, medical history and to evaluate the patient’s eligibility to use the iVR device. Radiologist reports and brain scans routinely acquired in clinical practice were extracted from patient’s medical files to extract the date of stroke, stroke etiology, presence of prior strokes and lesion side. In addition, the Barthel index which assesses independence in activities of daily living was extracted from the medical files.

To describe our patient sample and explore which patient characteristics may be associated with iVR feasibility, we administered a cognitive screen, and fatigue and mood scales. The Dutch Oxford Cognitive Screen (OCS-NL) (Demeyere, Riddoch, et al., 2015; Huygelier, Schraepen, Demeyere et al., 2019) was administered to screen for impairments in attention, memory, language, numeric cognition, and praxis. Age-adjusted norms were used to interpret test scores (Huygelier, Schraepen, Demeyere, et al., 2019). The Dutch Fatigue Severity Scale (FSS) (Nadarajah et al., 2017a; Rietberg et al., 2010) is a questionnaire containing nine questions by which the perceived severity of fatigue symptoms in the past week during daily-life situations are evaluated. The FSS shows good reliability and validity in stroke patients (Nadarajah et al., 2017b; Ozyemisci-Taskiran et al., 2019). The Hospital Anxiety and Depression Scale (HADS) (Zigmond & Snaith, 1983) measures core complaints of anxiety (7 items) and depression (7 items). The HADS addresses emotions felt in the past 4 weeks. We used the recommended cut-off score (≥8), which shows good sensitivity and specificity (Bjelland et al., 2002).

### 2.3. Procedure

This study consists of screening data collected in the first sessions of a clinical trial on the efficacy of an iVR neglect training (Huygelier et al., 2024b). Eligible stroke patients were invited for a cognitive screening. After providing written informed consent, stroke patients were enrolled in three screening sessions, which were planned according to the availability and cognitive abilities of each patient. In session 1, patients completed a demographic and medical questionnaire, the Dutch Oxford Cognitive Screen, the Weintraub Random Shape Cancellation test, the Behavioral Inattention Test (BIT) Star Cancellation test and the McIntosh Line Bisection test. In session 2, patients completed a Digital Cancellation test and in session 3, the Catherine Bergego Scale, the Fatigue Severity Scale and the Hospital Anxiety and Depression scale. Next, stroke patients who could safely use the iVR system were invited to participate in a second set of sessions: in session 4, they performed an iVR Tutorial and filled in the User Experience scale and the Simulator Sickness Questionnaire. In session 5, they performed the iVR Assessment and filled in the User Experience Scale again. Lastly, the Posner test was administered in session 6.

### 2.4. Statistical Analysis

Statistical analyses were performed in R (R Core Team (2023)). All scripts and data used in this study are accessible via https://osf.io/vuwjd/?view_only=fbb7eac1be994b329b0bfebafa996fed.

#### 2.4.1. Diagnostic accuracy

To investigate the diagnostic accuracy of the iVR assessment, a novel approach was developed (see Figure 1). Rather than classifying patients as having neglect or no neglect using conventional diagnostic cut-offs, we quantified the uncertainty regarding this classification and used this information to then estimate the diagnostic accuracy of the novel iVR test and contrast it with pen-and-paper and non-iVR digital tests.

**Figure 1.**
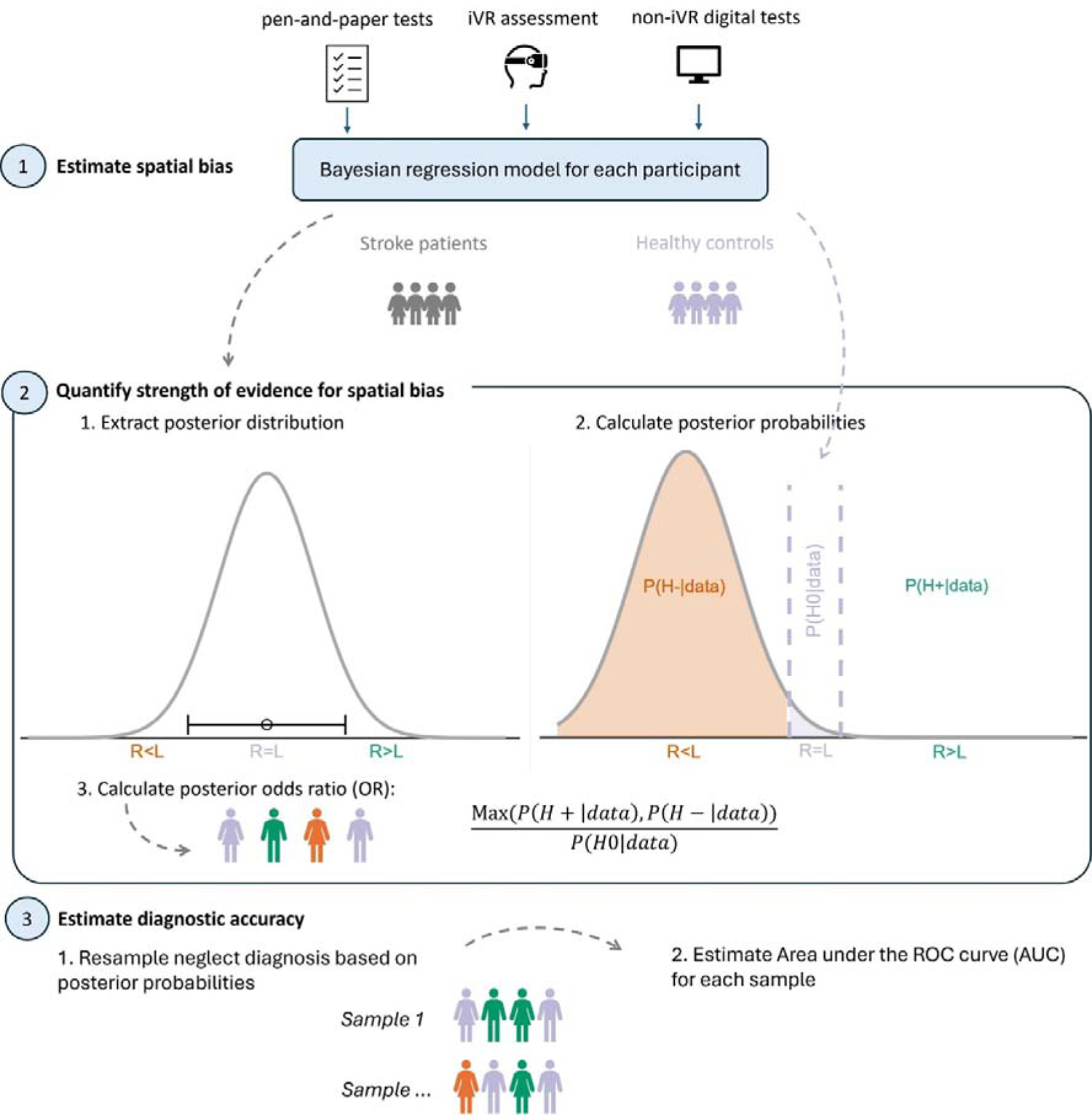
Overview of analysis approach. We first estimate the strength of evidence for or against a spatial bias per patient. Then, diagnostic accuracy is estimated, acknowledging the diagnostic uncertainty through resampling.

##### Step 1. Estimate a spatial bias per test

First, we estimated the spatial bias for each test using a Bayesian regression model (see Appendix A for details on the estimation and fit of the models) for each patient. The regression models enable to estimate the range of plausible spatial bias values for a patient, representing uncertainty in the measurements. The latter generates a data-driven interval estimate of spatial bias, rather than a point estimate. From these regression models, the posterior distribution (representing the range of plausible values for the spatial bias given the observed data) of an index of spatial bias for each test was extracted (see Appendix A for details).

##### Step 2. Quantify strength of evidence for or against a spatial bias

Then, based on the posterior distributions, the probability of three hypotheses given the data were derived: H0 = no spatial bias, H- = left-sided spatial bias (indicative of right-sided neglect) and H+ = right-sided spatial bias (indicative of left-sided neglect) (i.e., consistent with the Bayesian posterior probability). As healthy individuals may also show spatial biases different from the exact null value, we specified an interval of small spatial bias values that were considered equivalent to null (see Appendix A). For the digital tests, we used a range from the 5^th^ to the 95^th^ percentile of the spatial bias estimates in healthy controls (see Appendix A). For the pen-and-paper tests, we used the published cut-offs to determine the range (Cancellation tests: 10% difference (Huygelier, Schraepen, Demeyere, et al., 2020; Robertson et al., 1994), McIntosh Line Bisection: −.125 (right-sided neglect) and .075 (for left-sided neglect) (McIntosh et al., 2017)). We used a single normative range to interpret all patients’ scores, rather than a demographically adjusted range, as clinical cut-offs for spatial bias measures are typically not demographically corrected (e.g., McIntosh et al., 2005; Rengachary et al., 2009c) and as there was no evidence for an association of age and education with the absolute spatial biases in the digital tests (Appendix A).

As it is commonly recommended to diagnose neglect not merely on a single pen-and-paper test, but rather on a test battery, we integrated evidence for a spatial bias across the four pen-and-paper tests (i.e., OCS-NL Hearts Cancellation, Weintraub Random Shape Cancellation, BIT Star Cancellation and McIntosh Line Bisection). To integrate evidence across the four pen-and-paper tests, we averaged the three posterior probabilities across the tests. This allowed us to contrast the diagnosis based on a either a pen-and-paper test battery versus a single digital test. We also integrated evidence across all tests to obtain an idea of the most supported hypothesis across all tests. The latter was used as a reference to estimate the diagnostic accuracy. More information about how this novel integration method corresponds to traditional methods is reported in Appendix A.

To visualize results, we calculated a continuous index of evidence either in favor of a left-sided bias, no bias or a right-sided bias. To this end, we calculated the ratio of the posterior probability of a right-sided bias or a left-sided bias (taking the maximum of one of these) relative to the posterior probability of no spatial bias and added a sign. The resulting posterior odds ratios need to be interpreted as follows: a value above 1 favors the right-sided spatial bias hypothesis (H+) (i.e., left-sided neglect), and values smaller than −1 favor the left-sided spatial bias hypothesis (H-) (i.e., right-sided neglect). Values between −1 and +1 favor the no spatial bias hypothesis (H0). Absolute values above 3 or below .33 indicate moderate evidence, and absolute values above 10 or .10 indicate strong evidence for the corresponding hypothesis (Lee & Wagenmakers, 2014; Stefan et al., 2019). Absolute values in between 1 and 3 indicate weak evidence (i.e., inconclusive).

##### Step 3. Estimating and comparing the diagnostic accuracy

Last, to evaluate the diagnostic accuracy of the iVR assessment relative to the other tests (pen-and-paper and digital tests), we investigated the predictive strength of each test for the evidence for a spatial bias across all tests. To quantify how well each test predicts the other tests, we implemented Area Under the Receiver Operating Characteristic (ROC) curves. The area under the ROC curve (AUC) quantifies the test’s overall ability to discriminate between two groups. In this case, the AUC reflects how accurately a test can separate patients for which evidence favored H+ or not and patients for which evidence favored H- or not. The test with the highest AUC can be considered the most informative test for neglect diagnosis as this test is best able to predict outcomes on all administered tests. An AUC value of 1 indicates a perfect prediction, an AUC value of .5 indicates no predictive ability and an AUC value below .5 is worse than random guessing. To consider the uncertainty in diagnostic outcome, the neglect classifications (i.e., preferred hypotheses H+, H- or H0) were sampled from their respective posterior probabilities, generating 2500 samples. This way, we were able to obtain AUC confidence intervals which reflect the uncertainty in diagnostic accuracy. We then statistically contrasted the AUC’s between the iVR test and the other tests, by obtaining the 99% interval of the difference of the estimated AUC’s.

#### 2.4.2. Feasibility

To report on the representativeness of our patient sample, we calculated the completion rate of the iVR sessions and reported the main reasons for exclusion and drop-out. Furthermore, we compared the descriptive characteristics of two groups of patients: all patients who completed the iVR assessment (partially or fully) versus all patients who were recruited but who did not complete the iVR assessment. The latter can reveal whether certain patient characteristics were associated with iVR non-completion.

To evaluate cybersickness, we assessed the presence and severity of cybersickness symptoms based on SSQ ratings before versus after patients completed the iVR Tutorial with a Bayesian paired *t*-test. To evaluate user experience in iVR, we described how patients evaluated their experience in the iVR assessment based on their ratings on the User Experience scale.

Independent samples Bayesian *t*-tests (continuous variables) and Bayesian Chi-square tests (categorical variables) were used to assess differences between groups. The Bayes Factor (BF) quantifies the relative strength of evidence in favor of the alternative (H_1_) versus null hypothesis (H_0_). A BF_10_ > 3 is considered moderate-strength evidence in favor of the alternative hypothesis, while a BF_10_ < .33 is considered moderate-strength evidence in favor of the null hypothesis (Lee & Wagenmakers, 2014; Williams et al., 2017). All BFs in between .33 and 3 are considered inconclusive or weak evidence. For all BF’s, we used the default priors as implemented in the BayesFactor R package, which have been validated by Rouder et al. (2012).

## 3. Results

### 3.1. Participants

During the recruitment period (September 2021 – October 2023), we intended to assess 74 stroke patients with iVR. Of this sample, 52 patients fully completed and 2 patients partially completed the iVR assessment. Of the partially complete patients, one patient stopped the iVR assessment after 1.65 minutes (completing 22 trials) and another patient after 25.4 minutes (completing 160 trials), while the assessment took on average 27.05 minutes (225 trials) (*SD* = 4.94). Reasons for missing iVR data are reported in Figure 2. Patients completed all test sessions on average in 12 days (Mdn = 9.5, SD = 6.8, range: 2-45).

**Figure 2.**
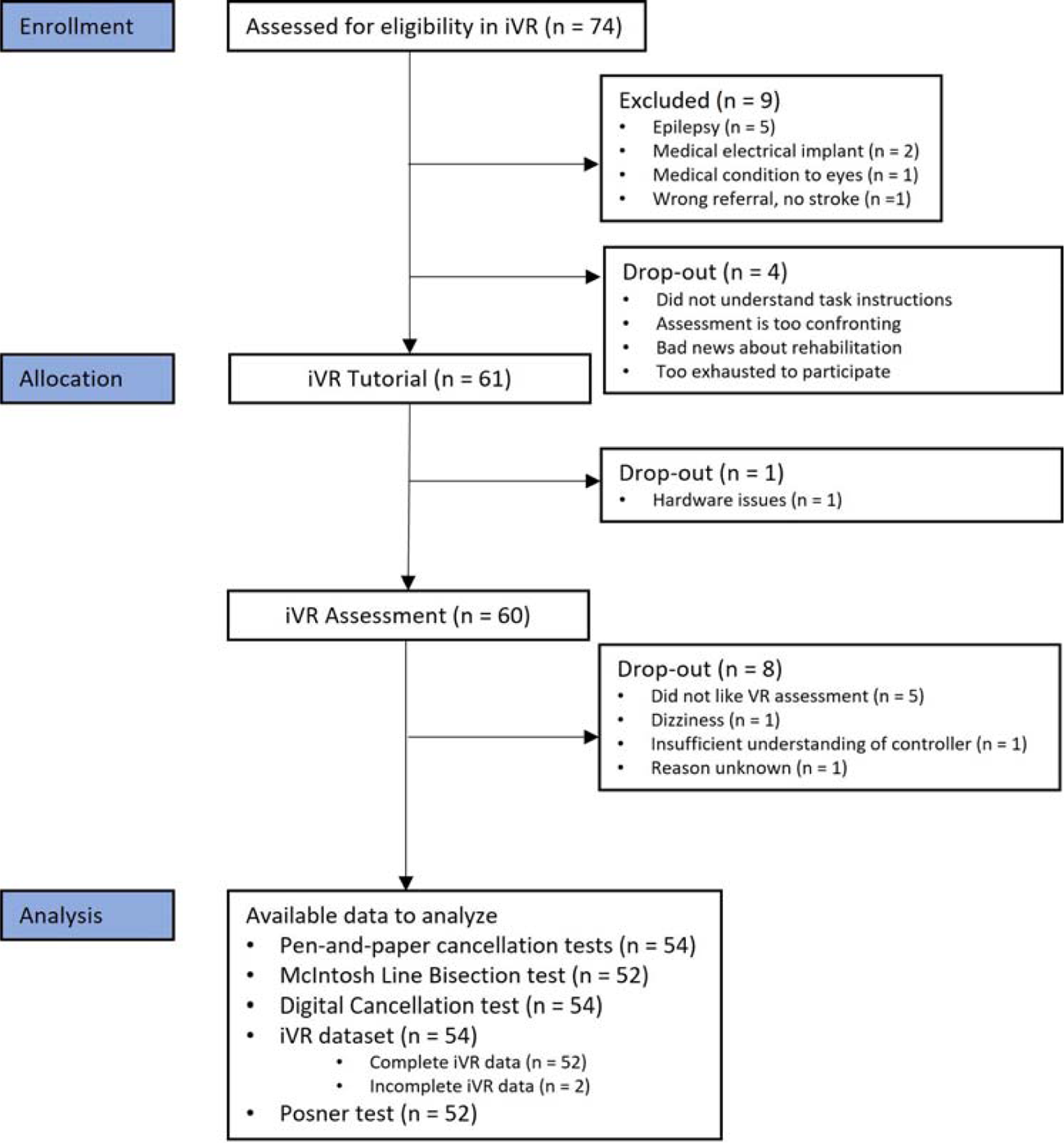
Flowchart of recruitment, enrollment and drop-out of the iVR sessions.

Patient descriptives of the two groups (patients who did versus who did not participate in iVR assessment) are reported in Table 3. In general, comparisons between patients who were tested versus not tested in the iVR assessment in demographic or clinical characteristics were either inconclusive or favored the absence of a difference (Table 3). There was one exception, namely there were significantly more first-ever stroke patients in the iVR-tested group (71%) than in the iVR-not-tested group (32%) (Table 3). Detailed descriptives of the patients who did not complete the iVR assessment are reported in Appendix B.

**Table 2.** Description of pen-and-paper neglect tests and their diagnostic accuracy.

| <u>Test</u> | <u>Test design</u> | <u>Sensitivity and Specificity</u> | <u>Comparison Test</u> | <u>References</u> |
| --- | --- | --- | --- | --- |
| OCS-NL Hearts Cancellation | Targets: 50<br>Distractors: 100 | Sensitivity = 65%<br>Specificity = 75% | Star Cancellation | Demeyere, Riddoch, et al., 2015 |
| Weintraub Random Shape Cancellation | Targets: 60<br>Distractors: 300 | Sensitivity = 51.7%<br>Specificity = / | CBS ($\geq 1$) | Weintraub & Mesulam, 1988; Kaufmann et al., 2020b |
| Star Cancellation test | Targets: 54<br>Distractors: 75 | Sensitivity = 50%<br>Specificity = / | CBS ($\geq 1$) | Halligan, Cockburn, & Wilson, 1991; Kaufmann et al., 2020b |
| McIntosh Line Bisection | Lines: 32 | Sensitivity = /<br>Specificity = / | / | McIntosh, 2017; McIntosh et al., 2005 |

**Table 3.** Patient characteristics of the intention-to-assess sample.

| Outcome | Assessed in VR<br>(n = 54) | Not assessed in VR<br>(n = 20) | $BF_{10}$ | Evidence |
| --- | --- | --- | --- | --- |
| | $M$ (SD) | $M$ (SD) | | |
| Age (years) | 59.37 (13.73) | 65.84 (13.55) | 0.99 | inconclusive |
| Gender (prop. males) | 0.63 | 0.47 | 0.73 | inconclusive |
| Education (years) | 13.8 (3.55) | 12.53 (2.65) | 0.59 | inconclusive |
| Time since stroke (days) | 55.57 (40.2) | 56.37 (25.7) | 0.27 | moderate for $H_0$ |
| First-ever stroke (prop.) | 0.71 (0.29) | 0.32 (0.68) | 30.96 | **strong for $H_1$** |
| Type of stroke (prop.) | 0.24 (HS),<br>0.76 (IS) | 0.32 (HS),<br>0.68 (IS) | 0.41 | inconclusive |
| Lesion side | 0.5 (R), 0.35 (L),<br>0.15 (BL) | 0.26 (R), 0.42 (L),<br>0.32 (BL) | 1.15 | inconclusive |
| Barthel index | 15.1 (5.74) | 13 (6.07) | 0.57 | inconclusive |
| CBS | 1.44 (2.28) | 0.75 (1.39) | 0.49 | inconclusive |
| Visual Field (OCS-NL) | 3.71 (0.72) | 3.74 (0.65) | 0.27 | moderate for $H_0$ |
| FSS | 3.85 (1.46) | 4.06 (1.4) | 0.31 | moderate for $H_0$ |
| HADS Anxiety | 6.52 (4.94) | 6.56 (4.21) | 0.28 | moderate for $H_0$ |
| HADS Depression | 5.96 (4.22) | 6.12 (3.98) | 0.29 | moderate for $H_0$ |

Note. HADS = Hospital Anxiety and Depression Scale: cut-off score $> 7$ (\*) = possible anxiety disorder or depression, cut-off score $> 10$ (\*\*) = likely anxiety disorder or depression. FSS = Fatigue Severity Scale: cut-off score $\geq 4$ = moderate to high fatigue. CBS = Catherine Bergego Scale: cut-off score $> 10$ = moderate neglect (\*), cut-off score $> 20$ = severe neglect (\*\*). Barthel Index: cut off score $< 10$ = severely in need of assistance, cut off score $< 5$ = completely dependent. R = right, L = left, BL = bilateral, lesions. HS = hemorrhagic stroke, IS = ischemic stroke. $BF_{10}$ = Bayes Factor. $H_0$ = null hypothesis. $H_1$ = alternative hypothesis.

### 3.2. Description of the diagnostic evidence for each test and correspondence with pen-and-paper test battery

#### Pen-and-paper tests

On the combined results of the four pen-and-paper tests, there was evidence in favor of a spatial bias for 7 of the 54 patients (13%, left-sided neglect: n = 6, right-sided neglect: n = 1). For these 7 patients, evidence was either weak (n=3), moderate (n=1) or strong (n=3) (Figure 3). For the remaining 47 patients (87%), evidence favored the no-bias hypothesis, with 10 patients (20%) in the weak evidence range.

**Figure 3.**
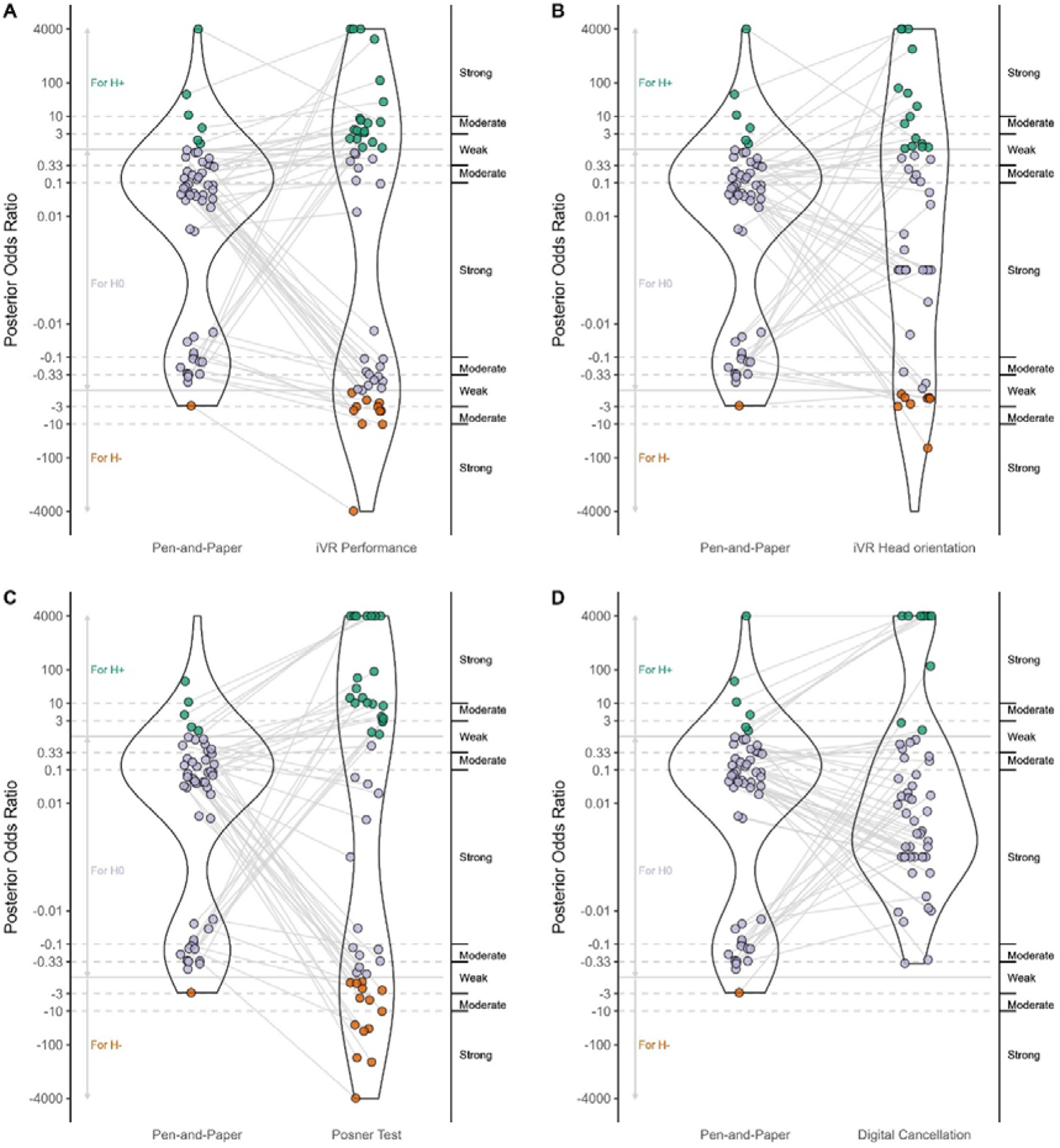
Posterior odds ratio for the pen-and-paper test battery and each digital outcome measure. H- = evidence in favor of a left-sided bias, H0 = evidence in favor of no bias, H+ = evidence in favor of a right-sided bias.

#### iVR performance

In general, evidence favored the spatial bias hypothesis for 33 of the 54 patients (61%), with 10 patients in the strong evidence range. iVR performance favored the spatial bias hypothesis for all patients (n = 7) for which the battery of pen-and-paper tests favored a spatial bias (Figure 3A). For patients for which pen-and-paper tests favored the no-bias hypothesis, 21 of the 47 patients (45%) remained in the no-bias category based on their iVR performance (Figure 3A). For the other 26 patients (55%), iVR performance indicated evidence in favor of a spatial bias (in contrast to the pen-and-paper tests). Within this group, evidence favoring a bias in iVR performance was strong for 6 patients (i.e., right-sided bias n = 4 and left-sided bias n = 2) and weak-to-moderate for 20 patients (i.e., right-sided bias n = 12 and left-sided bias n = 8) (Figure 3A).

#### iVR head orientation

In general, the spatial bias hypothesis was favored for 29 of the 54 patients (54%), with 14 patients in the strong evidence range. For iVR head orientation (Figure 3B), all patients (n = 7) with evidence favoring a spatial bias on the pen-and-paper test battery remained in the same category. For patients for which pen-and-paper tests favored the no-bias hypothesis, 25 of the 47 patients (53%) remained in the no-bias category on iVR head orientation (Figure 3B). For the other 22 patients (47%) evidence favored a bias in head orientation with strong evidence for 10 patients (i.e., right-sided bias n = 8 and left-sided bias n = 2) and weak-moderate evidence for 12 patients (i.e., right-sided bias n = 6 and left-sided bias n = 6).

#### Non-iVR digital Posner test

The Posner test indicated evidence in favor of the spatial bias hypothesis for 39 of 53 patients (74%) (Figure 3C), among which evidence was strong for 24 patients. 6 of the 7 patients with evidence favoring a spatial bias on the pen-and-paper test battery remained in the same category on the Posner test. Notably, 1 of the 7 cases changed from moderate evidence favoring a right-sided bias on the pen-and-paper tests to strong evidence favoring a left-sided bias on the Posner test (see Figure 3C). For patients for which pen-and-paper tests favored no bias, 14 of the 47 patients (30%) remained in the no-bias category while 32 patients (68%) were in the left- or right-sided bias category on the Posner test. For these patients, there was strong evidence in favor of a spatial bias for 18 patients (i.e., right-sided bias n = 12, left-sided bias n = 6) and weak-moderate evidence in favor of a bias for 13 patients (i.e., right-sided bias n = 6, left-sided bias n = 7).

#### Non-iVR Digital Cancellation test

On the Digital Cancellation test, there was evidence in favor of a spatial bias for 10 of the 54 patients (19%), among which evidence was strong for 8 patients (see Figure 3D). A total of 5 of the 7 patients with evidence favoring a spatial bias on the pen-and-paper tests remained in the same category, while evidence favored the no-bias hypothesis for 2 patients. For the 47 patients for which pen-and-paper tests favored the no-bias hypothesis, the Digital Cancellation test confirmed this for 42 patients (89%). Among the other 5 patients, evidence was always in favor of a right-sided bias with strong evidence for 3 patients.

### 3.3. Diagnostic Accuracy of iVR relative to other tests

For most patients (76%), test results (pen-and-paper battery, iVR and non-iVR digital tests) were not fully consistent. There were many patterns of agreements and disagreements among tests (see Figure 4).

**Figure 4.**
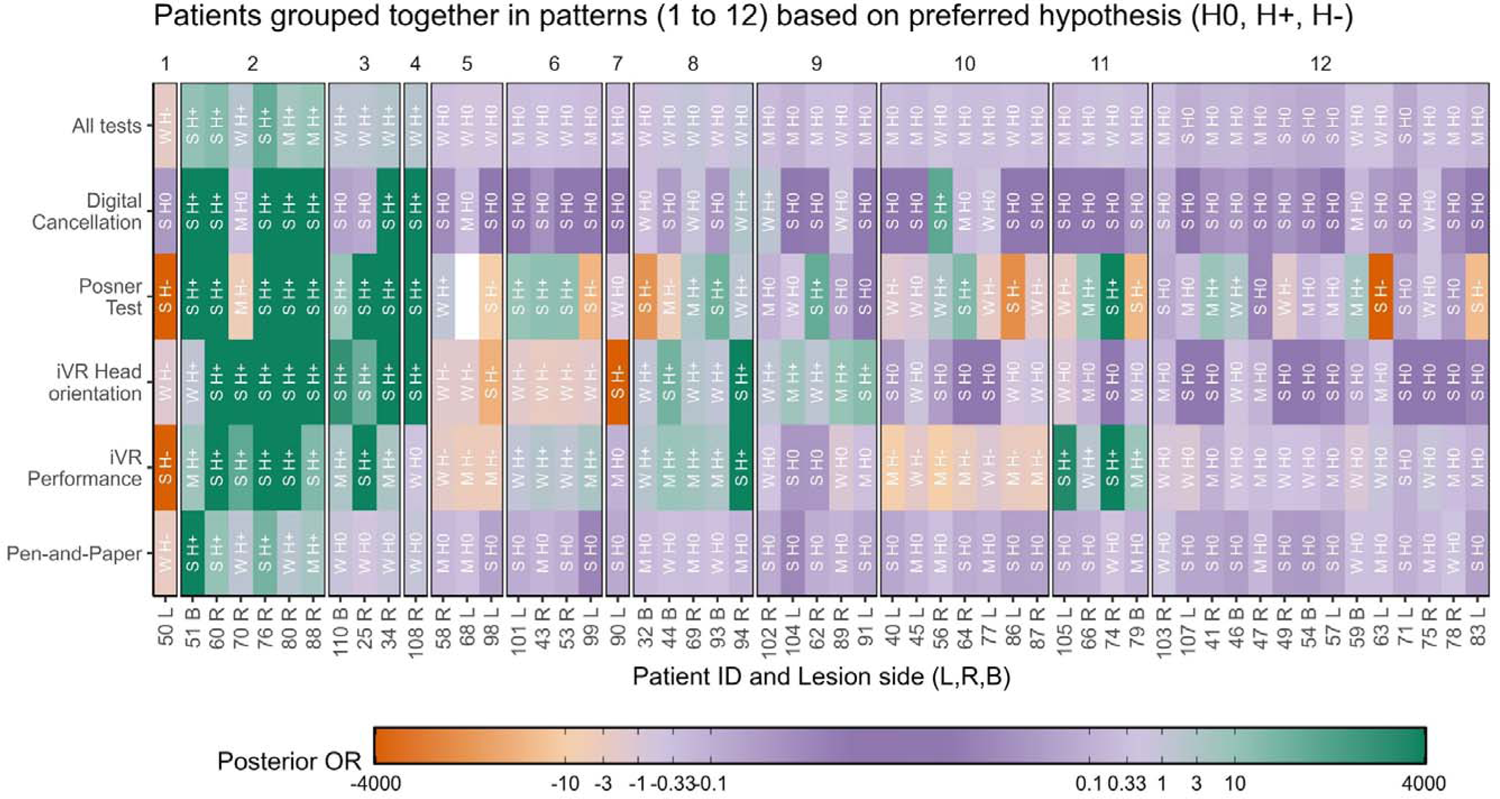
Posterior odds ratios for each patient on each digital test and the pen-and-paper battery. S = strong evidence, M = moderate evidence, W = weak evidence. H+ = right-sided bias, H0 = no bias, H- = left-sided bias. The colour represents the favoured hypothesis and the shade represents the strength of evidence (see Posterior OR legenda). Participants are grouped together according to their pattern (numbered from 1 to 12) of test results (i.e., which hypothesis was favoured across pen-and-paper, all tests and iVR outcomes). Posner data was not available (hence the white colour) for 68 due to drop-out.

That is, the first two groups depicted in Figure 4 (patterns 1 and 2) are patients for which pen-and-paper tests indicated evidence in favor of a spatial bias (Figure 4). iVR test results confirmed this for all patients. Next, there were 4 patients (patterns 3 and 4) for which pen-and-paper tests favored the no-bias hypothesis, while evidence across all tests contrasted this result. These patients all demonstrated a spatial bias on (one or both) iVR outcomes and the Posner test. The next patterns (patterns 5 – 12) consist of patients for which both the pen-and-paper tests and evidence integrated across all tests favored the no-bias hypothesis. Patterns 5 and 8 consist of patients that showed a consistent bias on iVR performance and iVR head orientation (8 patients). Patterns 6, 7, 9, 10 and 11 consist of patients for which iVR performance and iVR head orientation did not provide consistent results. Last, pattern 12 consists of patients for which the iVR test was consistent with the no-bias hypothesis on the pen-and-paper test battery and all tests combined. A few patients stand out. For instance, patient 94 had a spatial bias on each digital test, while the pen-and-paper tests favored the no-bias hypothesis. In addition, patient 74 showed a spatial bias on iVR performance and the Posner test, which was not confirmed by the other tests.

These results are consistent with the findings of the diagnostic accuracy (AUC). That is, to identify patients with a right-sided spatial bias (H+), the four digital tests had higher AUC values than the four pen-and-paper tests (Table 5, Figure 10A), but there was no statistically significant difference in the AUC values between the iVR outcomes and the other tests, when considering the uncertainty in the neglect status across all tests (see Appendix C, Table C1). In addition, the Posner and iVR outcomes had higher AUCs than other tests to identify left-sided spatial biases (H-), but again there was no statistically significant difference in AUCs between the iVR outcomes and other tests at an alpha of .01 (see Appendix C, Table C1).

**Figure 5.**
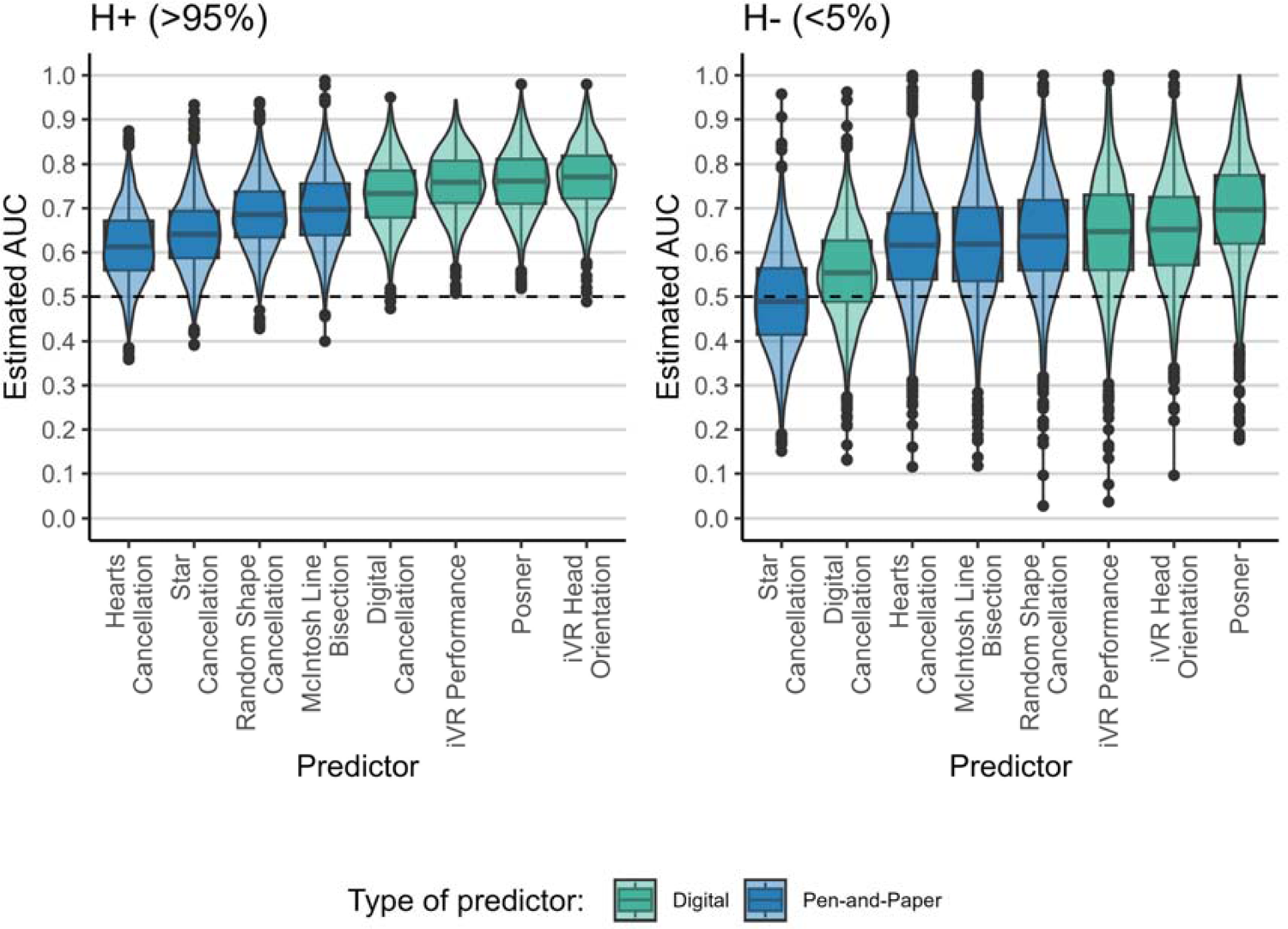
The estimated AUC value for the prediction of no bias (H0) versus right-sided bias (H+) (panel A) and left-sided bias (H-) (panel B) for all other outcomes per predictor (x-axis).

### 3.4. Cybersickness and user experience

#### Cybersickness

A total of 58 patients (including a subgroup of those that dropped out of the iVR sessions) completed the cybersickness questionnaire. We investigated whether there was evidence for a difference in cybersickness symptoms pre versus post iVR for each item using a Bayesian paired *t*-test (see Table 4). None of the items showed evidence for an increase in cybersickness symptoms pre versus post iVR (Table 4, Figure 6).

**Figure 6.**
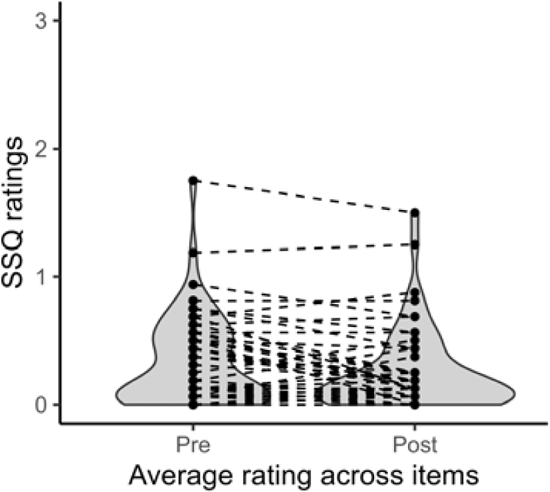
Average SSQ ratings across items.

**Table 4.**
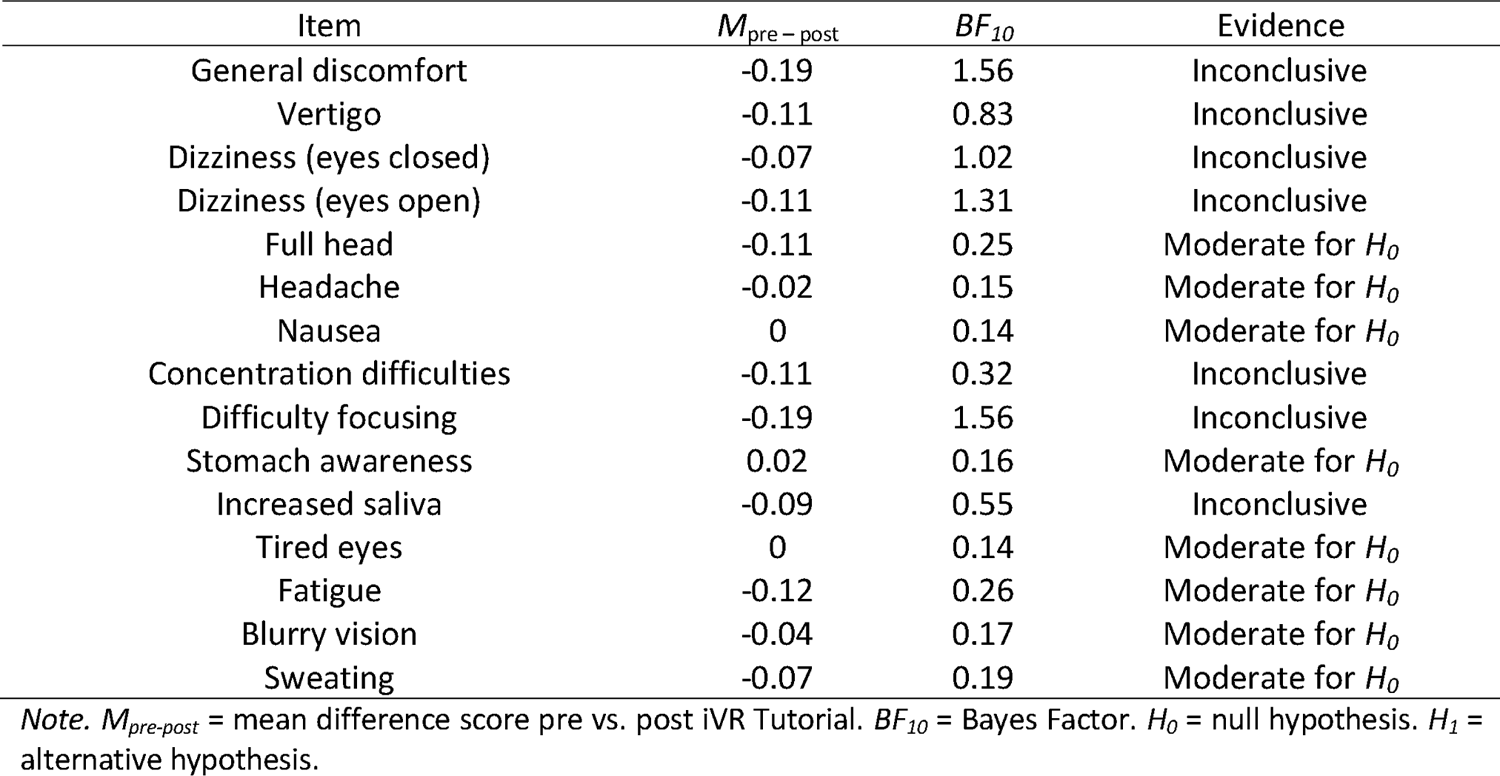
Results of cybersickness symptoms (SSQ items) pre vs. post iVR Tutorial.

**Table 5.** Estimates of best cut-off, AUC, sensitivity and specificity of each test to differentiate left- and right-sided biases from no bias in stroke patients (n = 54).

| Test | Cutoff | Right-sided bias (H+) |  |  | Cutoff | Left-sided bias (H-) |  |  |
| --- | --- | --- | --- | --- | --- | --- | --- | --- |
|  |  | AUC | Sensitivity | Specificity |  | AUC | Sensitivity | Specificity |
| Hearts Cancellation | .12 | .62 | .49 | .89 | -.05 | .61 | .70 | .63 |
| Star Cancellation | .08 | .64 | .47 | .92 | .02 | .49 | .78 | .42 |
| Random Shape Cancellation | .09 | .69 | .56 | .86 | -.03 | .64 | .68 | .66 |
| McIntosh Line Bisection | .07 | .70 | .59 | .87 | -.02 | .62 | .69 | .67 |
| Digital Cancellation | .09 | .73 | .60 | .88 | .03 | .56 | .84 | .45 |
| Posner test | .07 | .76 | .72 | .80 | -.02 | .69 | .74 | .72 |
| iVR performance | .05 | .76 | .70 | .77 | -.04 | .64 | .64 | .76 |
| iVR head orientation | 4.93 | .77 | .71 | .82 | -.71 | .65 | .74 | .65 |

#### User experience

A total of 56 patients (including a subgroup of those that dropped out of the iVR sessions) rated multiple items on the user experience scale to evaluate their experience during the iVR Tutorial and iVR Assessment. Most patients (80%) rated their user experience as positive (rating above 3 out of 5). On average, patients evaluated their Experience (M= 3.91, *SD* = 0.72) and the Usability positively (M=4.01,*SD* = 0.76), but their Presence was on average rated as 2.8 (*SD* = 0.85).

## 4. Discussion

In the current study, we investigated whether an iVR assessment that captures spatial biases in task performance (measured in head-contingent space) and head orientation (before trial-onset) could improve diagnostic accuracy for neglect. Crucially, we did not only compare the iVR assessment to conventional pen-and-paper tests, similar to previous studies (Belger et al., 2023; De Boi et al., 2024; Hougaard et al., 2021; Kim et al., 2021; Knobel et al., 2020; Painter et al., 2024; Thomasson et al., 2023), but we also compared it to commonly used non-iVR digital tests (i.e., the Posner test and Digital Cancellation test) that may be equally sensitive. To investigate diagnostic test accuracy, we used a new statistical approach that takes into account the ambiguity present in current diagnostic accuracy studies (i.e., when comparing novel tests to conventional tests that are no gold standard tests Azouvi et al., 2002; Huygelier, Moore, et al., 2020; Kohn et al., 2013). Using this approach, we investigated whether the iVR assessment could reduce the uncertainty of the neglect diagnosis, particularly in mild neglect cases that are difficult to detect with conventional tests.

Delving into the results, few patients showed significant and consistent spatial biases on the pen-and-paper tests (7 out of 54 patients). Each digital test confirmed the presence of a spatial bias for these cases. For these patients with severe spatial biases, digital tests had little added value to detect the presence of neglect. However, there was a separate group of patients (n = 6; 110, 25, 34,108,98 and 74 in Figure 4) for which pen-and-paper tests favored the absence of a spatial bias, while the Posner test and (one or both of) the iVR outcomes consistently indicated strong evidence for a spatial bias.

In contrast to the Posner and iVR assessment, the Digital Cancellation test did not favor evidence for spatial biases for most cases (only for 2 cases; 34 and 108 in Figure 4). Thus, merely presenting a conventional pen-and-paper Cancellation test in a digital format with more trials did not improve the sensitivity of the neglect assessment. In contrast, the increased number of trials enhanced the strength of evidence in favor of the *absence* of a spatial bias (as the increased number of Digital Cancellation trials allowed us to obtain more precise interval estimates). These results suggest that the Cancellation test is insufficiently sensitive to identify visuospatial impairments (as frequently reported, see for instance Azouvi et al., 2002; Bonato & Deouell, 2013; Checketts et al., 2021, 2021). These results clarify that this cannot be resolved by increasing the test’s reliability (Bailey et al., 2004), and provides further support for the necessity to use alternative paradigms than Cancellation tests to identify spatial neglect (Bonato et al., 2010).

Importantly, both the Posner test and iVR Assessment identified spatial biases that were left unnoticed by the pen-and-paper tests. Indeed, there is evidence that the Posner test is more sensitive than pen-and-paper tests in detecting (subtle) spatial biases, particularly in chronic stroke patients (Gillebert et al., 2011; Posner et al., 1984; Ptak & Bourgeois, 2024; Rengachary et al., 2009c; Vandenberghe et al., 2012). A crucial difference between the Cancellation tests (both pen-and-paper and digital format) and the Posner and iVR assessment is the use of sudden target onsets. Rather than searching for static targets in a sequential strategic way, patients must respond quickly to sudden target onsets in the Posner and iVR assessment at unpredictable locations (invalid cued targets in the Posner test and unknown target locations within the iVR assessment). This feature may be essential in obtaining a more sensitive neglect assessment. Additionally, this feature may increase ecological validity as problems in visuospatial attention for unpredictable and briefly presented (peripheral) targets likely has an extensive impact on visuospatial attention in daily life, such as safely participating in busy traffic (Spreij et al., 2020; van Kessel et al., 2010). The latter however requires empirical validation in future studies.

One of our main aims was to evaluate whether the iVR assessment had superior diagnostic accuracy relative to other digital tests. However, we found no evidence for superiority as the AUC values were highly similar between the Posner test and iVR assessment. Moreover, spatial biases on the Posner and iVR assessment were not always consistent. Several factors could contribute to these inconsistent test results, such as the distance of stimuli to the observer (Posner may be more sensitive to peri-personal neglect and the iVR test may be more sensitive to extra-personal neglect), task instructions (Posner test may represent covert attention more and the iVR test may represent overt attention more), stimulus eccentricities (Posner test may miss some patients for which problems are only clear in the periphery), and the outcome measures (recording reaction times in the iVR test may aid in identifying subtle spatial biases similar to the Posner test). Further research is needed to clarify whether the inconsistent test results are robust, and if so, which factors contribute to the inconsistencies. Spatial biases on iVR Performance and iVR Head Orientation were also not fully consistent. Here again, the two iVR measures might tap into distinct underlying attentional processes or may not be robust. That is, no explicit instructions were provided to participants regarding their head orientation, which introduced unintended variability in patients’ exploratory behavior. However, even with standardized instructions Hougaard et al. (2021) observed inconsistencies in the direction of biases of gaze (head and eye orientation combined) and biases on conventional neglect tests.

Apart from the diagnostic test accuracy of iVR compared to other neglect tests, we also evaluated the feasibility of the iVR assessment by assessing multiple parameters. Our results established that assessing neglect using iVR was feasible for the majority (70%) of our consecutively recruited left- and right-hemispheric stroke patients. Reasons for drop-out and descriptive characteristics of dropped-out patients varied, underlining that there is not a single profile of stroke patients for which iVR is unfeasible. Furthermore, self-reported cybersickness symptoms were low and did not change after performing the iVR Tutorial. Overall, user evaluation ratings of iVR were positive. To conclude, these feasibility results established that iVR is feasible in a representative group of the stroke population, further strengthening prior evidence of good feasibility of iVR neglect assessment (Table 1).

### 4.1. Conclusion

This study demonstrated that an iVR assessment and a Posner test detect spatial biases that were not picked up by conventional pen-and-paper tests and a Digital Cancellation test. Our new statistical approach allowed us to take into account an imperfect golden standard and allowed us to integrate evidence for neglect across tests in a principled way. However, we could not demonstrate significant superiority of the iVR assessment. This result can be attributed to the inconsistencies between tests, which highlight the complexity of neglect assessment. Future studies should further explore which test characteristics are crucial to obtain a precise and sensitive assessment of spatial neglect. Furthermore, this study established that the iVR assessment was feasible for the majority of stroke patients, with low cybersickness and positive user evaluations.

## Author contributions

Authors E.P., H.H. and C.R.G. designed the study. Authors E.P. and H.H. analyzed the data, made figures and tables and wrote the main manuscript text. Authors E.P. and H.H. reviewed the literature. Authors H.H., C.R.G and E.V. reviewed the manuscript. Authors E.P., N.T., K.M., H.H. and E.N. recruited participants and collected data.

## Data Availability

All preprocessed data produced are available online at https://osf.io/vuwjd/?view_only=fbb7eac1be994b329b0bfebafa996fed

## Acknowledgements

We would like to thank UZ Leuven Pellenberg for their contribution to this study. We would also like to thank Ayten Köksal, Jelle Loyaerts, Pauline Spiessens, Stien Vandeweyer and Maarten Spruyt for their contributions to the data collection.

## Research data for this article

Preprocessed data and analysis code can be found on https://osf.io/vuwjd/?view_only=fbb7eac1be994b329b0bfebafa996fed. Participants were assured raw data would remain confidential and would not be shared due to the sensitive nature of the data in this study.

---

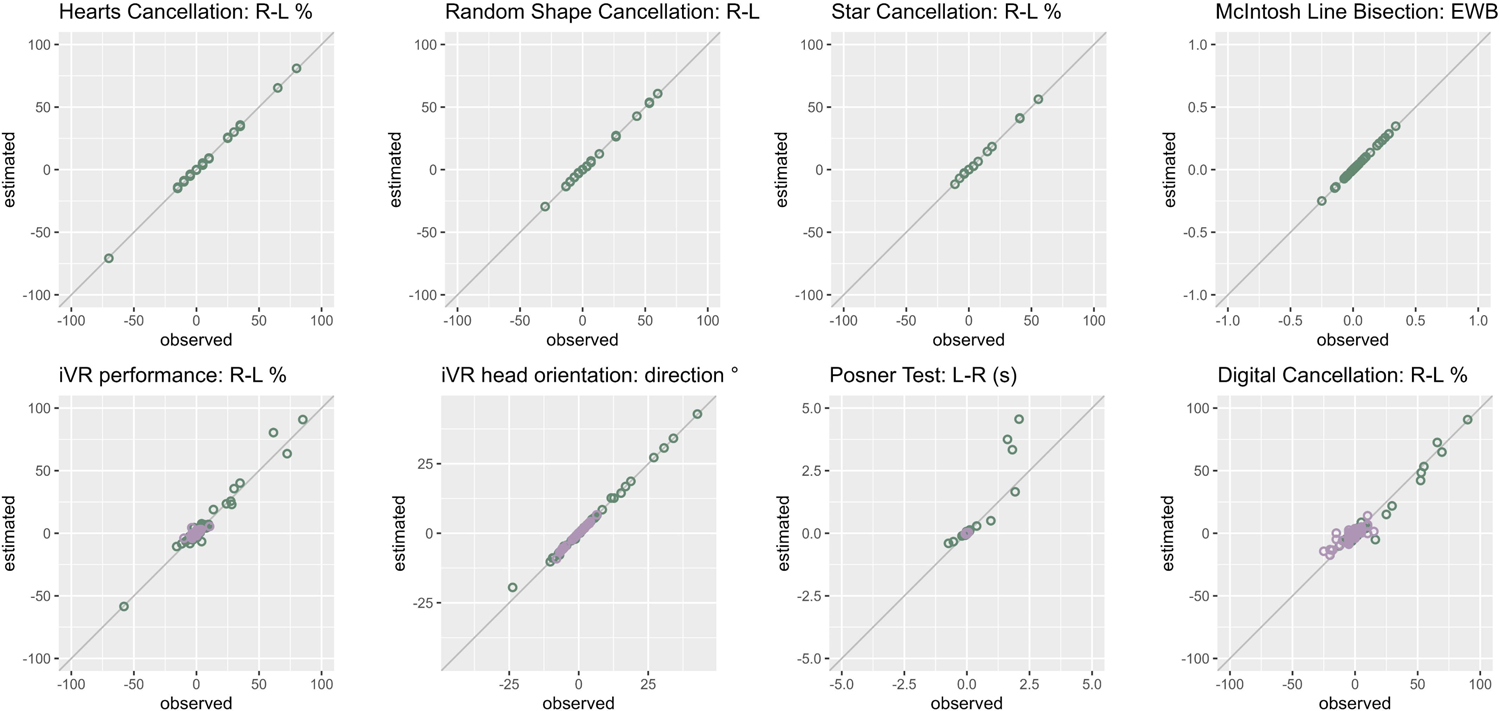

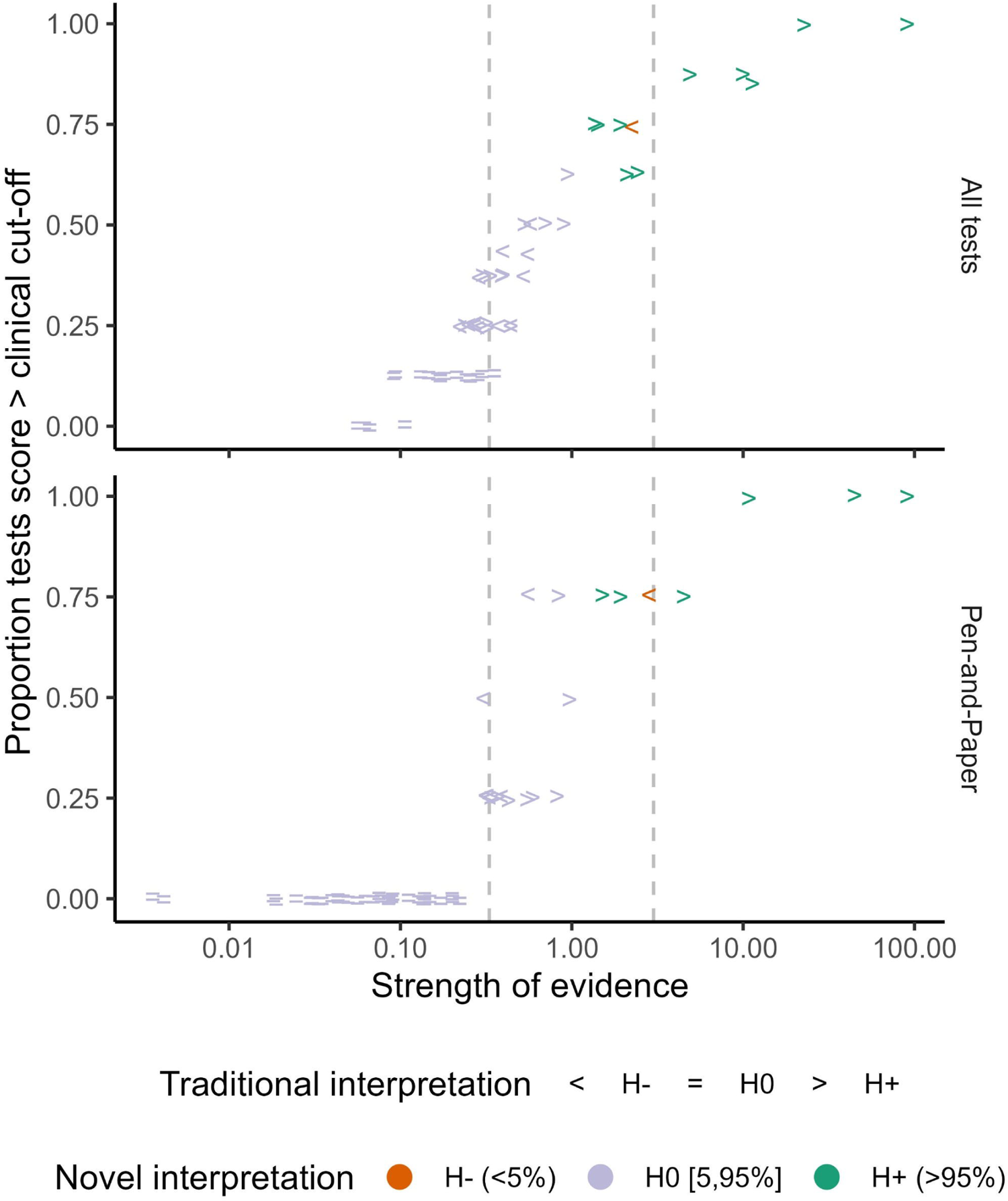

