## Appendices for "Does immersive VR improve diagnostic accuracy of post-stroke spatial neglect relative to conventional and digital tests?"

### Appendix A. Data-analysis additional information

Patient data: missing data and completion of tasks

A total of 54 patients completed or partially completed the iVR assessment. Of these 54 patients, not all non-iVR tests were completed or could be included in further analysis.

For the Posner test, 43 patients completed 400 trials, 5 patients completed 300 trials and 5 patients completed between 100 to 200 trials. One patient stopped participating during the iVR assessment and did not complete the Posner test afterwards. All patients who (partially) completed the Posner test complied with the task instructions. That is, the patients responded infrequently on catch trials (M = 2%, SD = 3%, range: 0 – 12%). Patients responded on average on 94% of the left-sided targets (SD = 19%, range: 0 – 100%) and on 99% of the right-sided targets (SD = 2%, range: 86 – 100%). For one patient, the bias in response times on the Posner test was impossible to estimate as this patient never responded on left-sided targets. Thus, spatial biases in response times on the Posner test could be estimated for 52 patients.

For the Digital Cancellation Test, 30 patients completed all 12 trials (each consisting of 50 targets and 100 distractors). The other 24 patients completed between 3 to 11 trials. All 54 patients were included in further analysis. For the McIntosh Line Bisection Test, two patients did not complete the test (1 due to comprehension problems and 1 due to an administration error). Thus, 52 patients were included in analyses. The three pen-and-paper cancellation tests were completed by all 54 patients.

#### Estimation of spatial biases

For each test, the spatial bias per participant was estimated with a regression model using the R brms v2.18.0 package (Bürkner, 2017).

For all cancellation tests and the iVR performance, a logistic regression model was used (Hosmer et al., 2013) to model the probability of detecting or cancelling a target depending on the spatial location of the target. For the pen-and-paper cancellation tests visual field (Left or Right) was the predictor. For the Digital Cancellation test, the horizontal position of the target (ranging from -13 cm to +13 cm) was used as a predictor. For the iVR performance, the horizontal position of the target (ranging from -30° to 30°) was used as a predictor. To analyze the response times on the Posner test, a censored shifted log-normal regression model was used (Rouder, 2005) modelling the response times as a function of the target location (i.e., Left or Right). This model is suitable for analysing response times when a response time limit (i.e., censoring) is implemented. For the Line Bisection test a linear regression model was used to model the response position as a function of the side of the endpoint (i.e., left or right) and the distance of the endpoint to the page midline (i.e., 4 or 8 cm). For iVR head orientation, an intercept model with a distribution of the Von Mises family was used (Damien & Walker, 1999). The Von Mises distribution is analogous to the normal distribution but for circular data (i.e., direction of an angle) (Damien & Walker, 1999).

Based on the regression coefficients, an estimate of the spatial bias was inferred. For all pen-and-paper cancellation tests estimates of the left-right difference in the proportion cancelled were inferred. For the iVR assessment, the difference in proportion detected targets for left-sided or right-sided targets at 25° eccentricity was estimated (i.e., iVR performance). For the Digital Cancellation test, the difference in cancelled targets at 12 cm eccentricity was estimated (close to the boundaries of the page). For the Posner test, estimates of the left-right difference in median response times (seconds) were derived. For the Line Bisection test, estimates of the normalized endpoint weighting bias were derived following McIntosh et al. (2017). For iVR head orientation estimates in degrees (0° = straight ahead, < 0° = left, > 0° = right) were derived.

Full model specifications are available in the analysis code, which is accessible via https://osf.io/vuwjd/?view_only=fbb7eac1be994b329b0bfebafa996fed. For all Bayesian models, we used the default (weakly informative) priors provided by the R brms package as this resulted in good model fit and model convergence. All models were fitted using Markov Chain Monte Carlo (MCMC) sampling. For each model, 4 independent chains were ran in parallel, using 4 processor cores. Each chain performed 2000 iterations, resulting in a total of 8000 samples per model. The first 1000 iterations were discarded as warm-up, leaving 1000 post-warm-up samples per chain for inference.

#### Quality checks of the regression models

We assessed model convergence using the potential scale reduction factor (R-hat) and effective sample size. All R-hat values were lower than 1.05 indicating good model convergence. In addition, we checked the recovery of the observed spatial biases for each test by comparing the observed with the estimated biases inferred from the models (Figure A1). For all pen-and-paper tests, the observed and estimated spatial biases were highly associated (Figure A1). For iVR performance, recovery was also good, but there was more variation. This is due to the fact that we estimate the bias at an eccentricity of 25°, but we calculated the observed bias for target eccentricities from 20 to 30° (to have enough datapoints). A similar logic applies to the digital cancellation test. For the Posner test the recovery is good, but one can see that extreme biases do not align perfectly. This is not a signal of bad model fit, but rather the effect of including the censoring into the regression model. That is, observed bias in response times is an underestimation of the true bias one would have observed if the response time limit would have been larger. The regression model corrects for this response time limit considering the percentage missed trials. As the spatial bias estimates and observed values corresponded well to each other (Figure A1), all further analysis were performed on the spatial bias estimates.

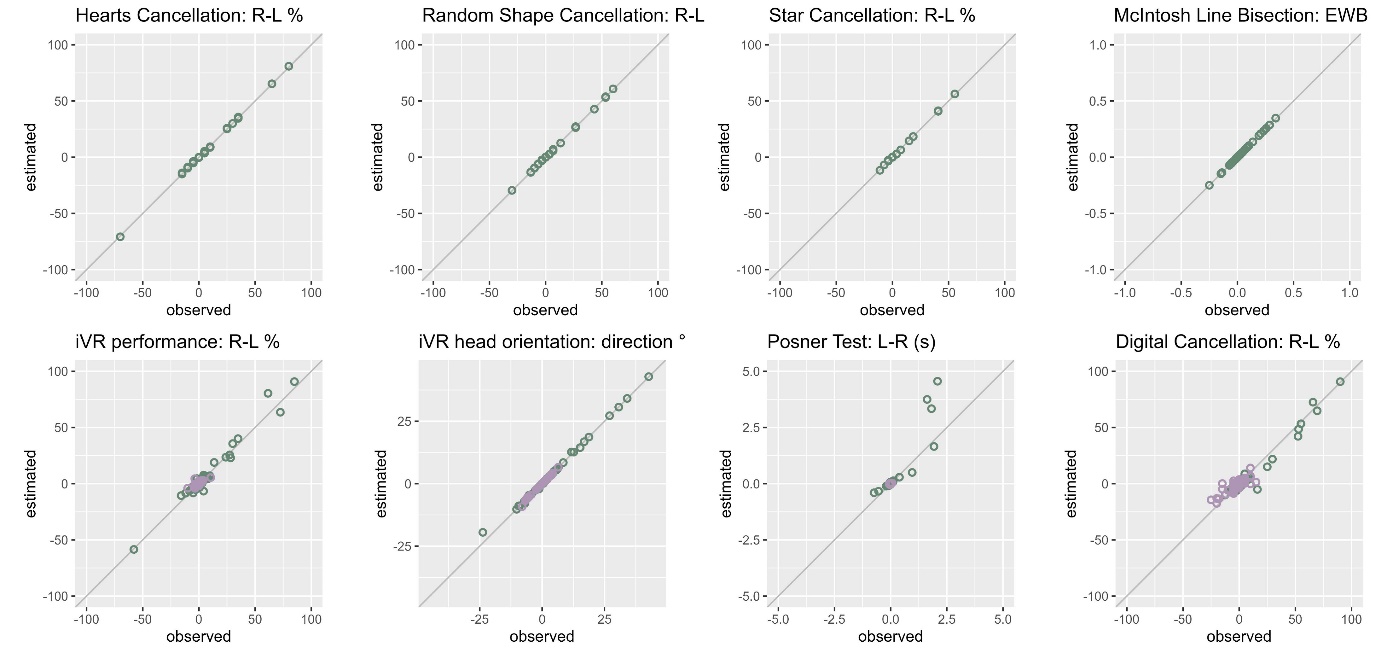

Figure A1. The association of the observed spatial bias per test and the estimated spatial bias using the regression models. The green dots are the estimates for the patients (n=54) and the purple dots for the healthy controls (n = 56).

#### Establishing normative data for the digital tests

##### Introduction

We planned to use fixed cut-offs to interpret spatial biases rather than demographically corrected cut-offs, as this is common in neglect research and as prior studies demonstrated no association of age or education with spatial biases in cancellation tasks (Benjamins et al., 2018). However, as our digital tests are novel, we assessed whether this absence of an effect is replicated in our sample of healthy participants. We thus assessed whether spatial biases in the digital tests (i.e., Digital Cancellation test, iVR assessment and the Posner test) were associated with age and education. Given the prior studies, we predicted that age and education would not be associated with spatial biases in the tasks employed in this study.

##### Method

###### Participants

Healthy controls were recruited via a database of elderly that have participated in previous studies of the research group. Healthy controls had to have normal or corrected-to-normal vision and hearing, no history of neurological or severe psychiatric disorders and no medical electrical implant or epilepsy.

###### Procedure

Healthy controls completed two 1-hour test sessions. We first administered a semi-structured interview to obtain demographic information and then screened for mild cognitive impairment using the Dutch version of the Montreal Cognitive Assessment (MoCA) (Nasreddine et al., 2005). Then, the Digital Cancellation test was administered. In session 2, healthy controls performed the Posner test, iVR Tutorial and iVR Assessment.

###### Data-analysis

We fitted two lognormal regression models on the absolute spatial bias for four outcome measures: absolute difference in the percentage cancelled targets between the left and right visual field (Cancellation test), absolute difference in response times between the left and right visual field (Posner test), absolute median head orientation in degrees and absolute difference in percentage detected targets for the iVR task. The first regression model (M0) was an intercept-only model (assuming a constant average and standard deviation), while the second model (M1) included the main effects of age, years of education and their pairwise interaction (i.e., demographic model).

All models were fit using 4 chains, 4000 iterations per chain and default priors in brms. All models converged with R-hat values lower than 1.05 and the graphical posterior checks indicated good recovery of the observed data (mean, median, standard deviation, minimum and maximum).

Then, the predictive performance of the intercept-only model and demographic model were estimated and compared using Leave-One-Out Cross-Validation (Sivula et al., 2023). The index for model fit is the Expected Log-Predictive Density (ELPD). Differences in ELPD larger than 4 suggest a difference in predictive performance of the two models (Sivula et al., 2023).

##### Results

###### Participants

A total of 56 healthy controls completed the digital tests (i.e., computerized cancellation, Posner test, VR test). They had an average age of 65.98 years (Mdn = 69, SD = 13.64, 24 – 85) and 15.5 years of education (Mdn = 15, SD = 3.22, 10-25). A total of 30 healthy controls were female (55%) and 49 were right-handed (88%). None of the healthy controls performed below the MoCA cut-off score of 26 (Nasreddine et al., 2005).

###### Descriptive of task performance

All Posner data of healthy controls were included in analysis as they performed well on the Posner test, responding infrequently on catch trials (M = 0%, SD = 1, range: 0 – 3%) and almost always on target-trials (Left: M = 100%, SD = 0, range: 99 - 100; Right: M = 100%, SD = 0, range: 99 - 100). One healthy control completed 3 trial blocks of the Posner test, while all other participants completed all 4 trial blocks. All other tests were fully completed by the healthy controls. Descriptives of the spatial biases and absolute biases for all healthy controls are reported in Table A1.

| Table A1. Descriptives of spatial biases and absolute biases in healthy controls (n = 56). | | | | | | | | |
| --- | --- | --- | --- | --- | --- | --- | --- | --- |
| Test | Outcome | M | Mdn | SD | Min | Max | 5^th^ pc | 95^th^ pc |
| Cancellation Test | R-L % | -1 | 0 | 6 | -18 | 14 | -13 | 5 |
|  | \|R-L\| % | 4 | 3 | 4 | 0 | 18 | .00 | 14 |
| Posner test | L-R ms | 0 | 0 | 20 | -40 | 30 | -23 | 30 |
|  | \|L-R\| ms | 10 | 10 | 10 | 0 | 40 | .00 | 31 |
| iVR Performance | R-L % | 0 | 0 | 2 | -4 | 5 | -3 | 3 |
|  | \|R-L\| % | 1 | 1 | 1 | 0 | 5 | .00 | 4 |
| iVR head orientation | Direction ° | -.27 | .4 | 3.1 | -9.2 | 6.6 | -6.3 | 3.6 |
|  | \|Direction\| ° | 2.3 | 1.7 | 2.1 | .04 | 9.2 | 0 | 6.5 |

###### Association of absolute spatial bias with age and education

For each outcome measure, the best-fitting model was the intercept-only model (M0) (Table A2). Indeed, the 95% credible intervals of the coefficients for age and education did not exclude zero as a plausible value for each spatial bias outcome (Table A3).

| Table A2. Model comparison for the digital outcome measures. | | | | |
| --- | --- | --- | --- | --- |
| Test | ELPD M0 | ELPD M1 | ELPD  M1 – M0 | SE ELPD  M1 – M0 |
| Cancellation Test | 109.3 | 107.3 | -2.0 | 2.5 |
| Posner Test | 176.0 | 174.0 | -1.9 | 7.0 |
| iVR Performance | 348.3 | 346.5 | -1.8 | 2.5 |
| iVR Head Orientation | -105.8 | -108.9 | -3.1 | 0.6 |
| Table Note. ELPD = Expected Log-Predictive Density, M0 = intercept-only model, M1 = model including age and education, SE = standard error of the ELPD. | | | | |

| Table A3. Estimated point values and 95% credible intervals for the demographic models (M1). | | | | | |
| --- | --- | --- | --- | --- | --- |
| Test | Intercept | Age | Education | Age * Education | Sigma |
| Cancellation Test | -.96  [-52.09, 48.05] | -.10  [-.80, .63] | -.55  [-3.33,2.36] | .01  [-.03, .05] | 4.90  [4.05, 5.95] |
| Posner Test | -8.85  [-21.72, 4.19] | .07  [-.11, .26] | .21  [-.53, .96] | -.00  [-.01,.01] | 1.29  [1.07, 1.57] |
| iVR Performance | -3.41  [-108.37, 96.40] | .08  [-1.35, 1.58] | -.14  [-5.83, 5.81] | -.01  [-.09,.07] | 9.76  [8.11, 11.86] |
| iVR Head Orientation | -.69  [-11.99, 11.45] | .01  [-.16, .18] | .07  [-.62, .72] | -.00  [-.01, .01] | 1.18  [.98, 1.45] |

##### Conclusion

There was no evidence for an association between absolute spatial biases on the four digital outcomes and age and education. As age and education were not significantly associated with spatial biases and as it is common to use fixed cut-offs to interpret spatial biases (rather than demographically corrected cut-offs) we created fixed cut-offs for the digital tests using the 5^th^ and 95^th^ percentiles of the spatial biases in the healthy controls.

#### Comparison between evidence integration: novel and traditional methods

##### Introduction

When administering a test battery to diagnose neglect it is common to set a threshold on the number of positive tests (tests on which a patient scores above a normative cut-off) required for diagnosis.

##### Method

To contrast the traditional method of integrating results across a test battery to our novel approach, we employed a criterion of at least 25% positive tests (i.e., ¼ positive pen-and-paper tests or 2/8 positive tests). This criterion corresponds to criteria used in prior studies, which employed criteria ranging from a liberal 17% to a more conservative 67% positive tests (Huygelier et al., 2020).

##### Results

According to these criteria, on the set of 4 pen-and-paper tests, 21 of the 54 patients (39%) were considered to have a spatial bias (i.e., have at least one positive test score). On the set of all 8 tests, 36 of the 54 patients (67%) were considered to have a spatial bias (i.e., have at least two positive test scores). In contrast, when diagnosis was based on the posterior evidence across the 4 pen-and-paper tests, only 7 of the 54 patients (13%) were considered to have a spatial bias. On the set of all 8 tests, 11 of the 54 patients (20%) were identified with a spatial bias.

When comparing the traditional method of interpreting scores on the test battery to the posterior evidence for the pen-and-paper test battery, we can see that a criterion of ¼ positive tests is liberal as it corresponds to a criterion of .30 for the posterior odds ratio. That is, all patients with a posterior odds ratio > .30 are classified as having a spatial bias according to the traditional method using a test battery of 4 pen-and-paper tests. For the criterion of 2/8 positive tests, this corresponded to a posterior odds ratio > .22, which is even more liberal. That is, all posterior odds ratio’s lower than 1 represent evidence in favor of the no-bias hypothesis and all posterior odds ratios lower than .33 are considered moderate evidence in favor of the no-bias hypothesis.

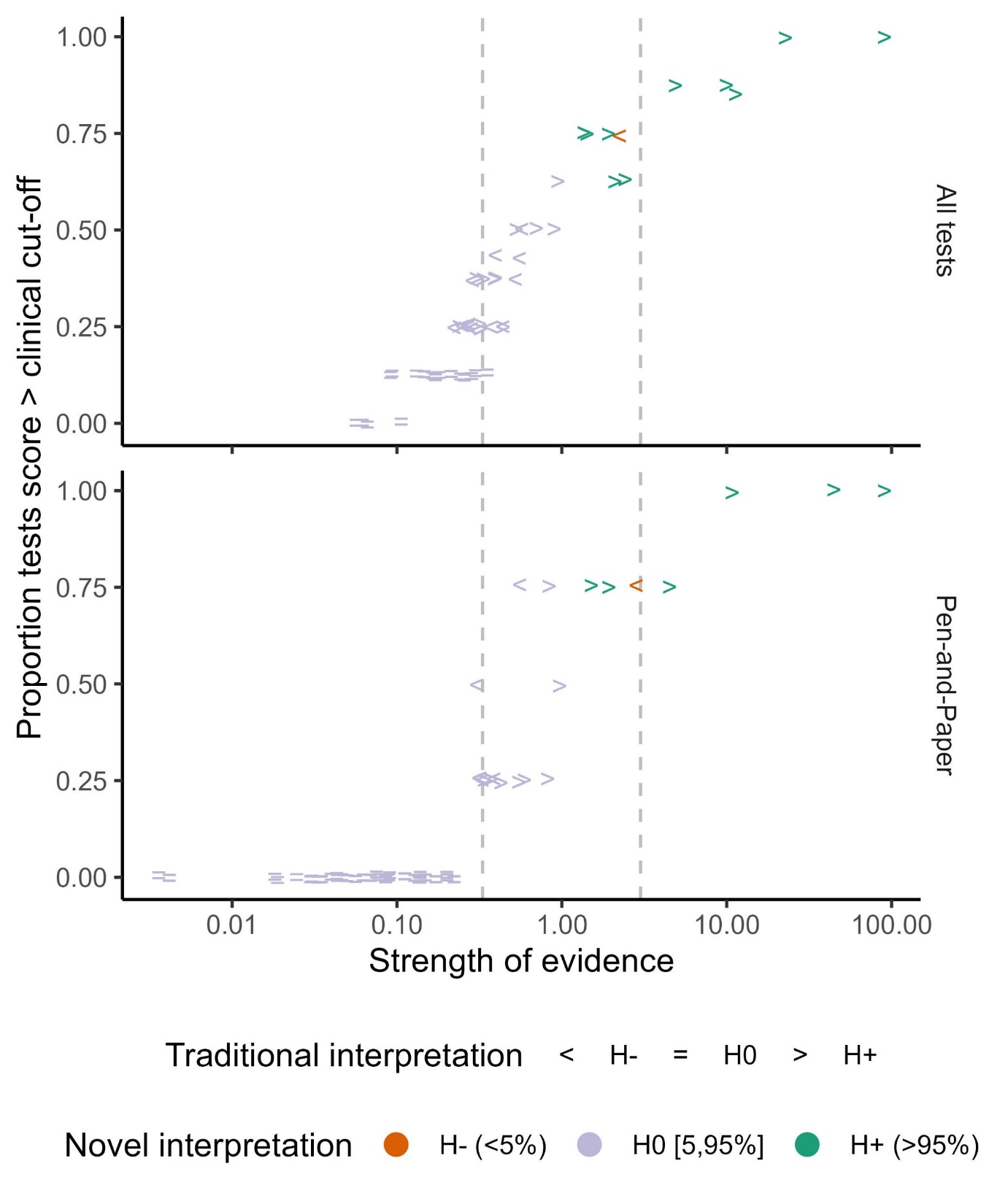

Figure A2. Correspondence between traditional method and novel method to identify a spatial bias for all tests and pen-and-paper tests. The strength of evidence in favour or against a spatial bias is depicted on the x-axis, while the proportion of positive tests is depicted on the y-axis. The two dashed lines demarcate the range of weak evidence going from 0.33 to 3. Evidence above 3 is interpreted as moderate evidence in favour of a bias. Evidence below .33 is interpreted as moderate evidence in favour of no bias.

### Appendix B. Patient descriptives of all patients who did not complete the iVR test

|  | Complete iVR data (n = 52) | Incomplete iVR data (n = 2) | | No iVR data (n = 8) | | | | | | | |
| --- | --- | --- | --- | --- | --- | --- | --- | --- | --- | --- | --- |
| Outcome | *M* (*SD*) | VR051 | VR068 | VR055 | VR065 | VR067 | VR073 | VR082 | VR092 | VR095 | VR097 |
| Age (years) | 58.46 (13.13 | 87 | 79 | 75 | 81 | 82 | 71 | 45 | 64 | 66 | 63 |
| Gender (prop. males) | 0.63 | M | M | F | F | F | M | F | M | F | M |
| Education (years) | 13.67 (3.47) | NA | 20 | 9 | 16 | NA | 11 | 12 | 12 | 15 | 11 |
| Time since stroke (days) | 55.46 (40.97) | 61 | 56 | 37 | 43 | 37 | 51 | 54 | 71 | 42 | 46 |
| Lesion side | 0.52 (R), 0.35 (L), 0.13 (BL) | BL | L | BL | BL | L | L | BL | L | R | BL |
| Reason for drop-out in iVR | -- | Insufficient understanding of controller | Reason unknown due to language problems | Does not like VR | Dizziness | Does not like VR | Does not like VR | Hardware issues | Does not like VR | Does not like VR | Does not like VR |
| Time spent in VR (minutes) | 27.05 (4.94) | 1.65 | 25.38 | -- | -- | -- | -- | -- | -- | -- | -- |
| Cybersickness (pre) | 0.33 (0.21) | 0.5 | 0.44 | NA | NA | 0.63 | NA | NA | NA | 0.25 | 0.31 |
| Cybersickness (post) | 0.25 (0.16) | 0.25 | 0.13 | NA | NA | 0.69 | NA | NA | NA | 0.5 | 0.19 |
| User experience | 3.51 (0.64) | NA | 2.17 | NA | NA | 2.78 | NA | NA | NA | 2.57 | 2.65 |
| Controllers were easy to use | 3.47 (1.26) | NA | 2 | NA | NA | 1 | NA | NA | NA | 2 | 2 |
| Barthel index | 15.42 (5.6) | 9 | 5 | 1 | 12 | 14 | 6 | 20 | 13 | 15 | 16 |
| CBS | 1.38 (2.3) | 3 | 3 | 1 | 0 | 0 | 4 | 0 | 0 | 0 | 2 |
| Visual Field (OCS-NL) | 3.71 (0.73) | NA | 4 | 4 | 4 | 2 | 4 | 4 | 4 | 4 | 4 |
| FSS | 3.83 (1.48) | 3.44 | 5.11 | 6 | 4.89 | 5.44 | 4.78 | 5 | 3.11 | 4 | 3.11 |
| HADS Anxiety | 6.75 (4.89) | 0 | 1 | 12 | 8 | 10 | 9 | 10 | 5 | 8 | 1 |
| HADS Depression | 6.1 (4.23) | 4 | 1 | 7 | 4 | 14 | 7 | 8 | 5 | 7 | 1 |

Table B1. Characteristics of patients who completed vs. dropped out during iVR sessions

*Note*. HADS = Hospital Anxiety and Depression Scale: cut-off score > 7 (*) = possible anxiety disorder or depression, cut-off score > 10 (**) = likely anxiety disorder or depression. FSS = Fatigue Severity Scale: cut-off score ≥ 4 = moderate to high fatigue. CBS = Catherina Bergego Scale: cut-off score > 10 = moderate neglect (*), cut-off score > 20 = severe neglect (**). Barthel Index: cut off score < 10 = severely in need of assistance, cut off score < 5 = completely dependent. OCS-NL = Dutch Oxford Cognitive Screen. R = right, L = left, BL = bilateral, lesions. Cybersickness ratings ranged from 0 (= no symptoms) up until 3 (= severe symptoms) . User experience and controller ratings ranged from 0 (= negative rating) up until 5 (= positive rating).

### Appendix C.

| Table C1. Estimated differences in AUCs between the iVR outcome measures and all other tests. | | | | | |
| --- | --- | --- | --- | --- | --- |
| iVR  outcome  measure | Spatial  bias  direction | Reference  Test | Difference in AUC | | |
|  |  |  | Mdn | 99% CI | |
| iVR  Head  Orientation | H- (<5%) | Digital Cancellation | 0.09 | -0.28 | 0.46 |
|  |  | Hearts Cancellation | 0.03 | -0.35 | 0.41 |
|  |  | McIntosh Line Bisection | 0.03 | -0.38 | 0.43 |
|  |  | Posner | -0.05 | -0.43 | 0.35 |
|  |  | Random Shape Cancellation | 0.01 | -0.38 | 0.40 |
|  |  | Star Cancellation | 0.16 | -0.21 | 0.52 |
|  | H+ (>95%) | Digital Cancellation | 0.04 | -0.20 | 0.28 |
|  |  | Hearts Cancellation | 0.15 | -0.10 | 0.40 |
|  |  | McIntosh Line Bisection | 0.07 | -0.19 | 0.33 |
|  |  | Posner | 0.01 | -0.24 | 0.25 |
|  |  | Random Shape Cancellation | 0.08 | -0.16 | 0.31 |
|  |  | Star Cancellation | 0.13 | -0.14 | 0.37 |
| iVR  Performance | H- (<5%) | Digital Cancellation | 0.09 | -0.31 | 0.49 |
|  |  | Hearts Cancellation | 0.03 | -0.36 | 0.44 |
|  |  | McIntosh Line Bisection | 0.03 | -0.42 | 0.43 |
|  |  | Posner | -0.05 | -0.45 | 0.40 |
|  |  | Random Shape Cancellation | 0.01 | -0.38 | 0.42 |
|  |  | Star Cancellation | 0.16 | -0.25 | 0.54 |
|  | H+ (>95%) | Digital Cancellation | 0.03 | -0.22 | 0.26 |
|  |  | Hearts Cancellation | 0.14 | -0.11 | 0.38 |
|  |  | McIntosh Line Bisection | 0.06 | -0.22 | 0.30 |
|  |  | Posner | 0.00 | -0.24 | 0.23 |
|  |  | Random Shape Cancellation | 0.07 | -0.17 | 0.31 |
|  |  | Star Cancellation | 0.12 | -0.14 | 0.35 |
